# SchistoTrackVideoNet: multilabel deep learning-based classification of schistosomal periportal fibrosis from ultrasound video

**DOI:** 10.64898/2026.06.01.26354613

**Authors:** Eloise Ockenden, Victor Anguajibi, Simon Mpooya, Benjamin Ntegeka, Tymothy Mugume, Betty Nabatte, Narcis Kabatereine, J. Alison Noble, Goylette F. Chami

## Abstract

Schistosomiasis causes a complex, difficult to diagnose form of liver fibrosis with high rates of life-threatening morbidity in resource-poor settings where there are often no trained sonographers. Protocols for diagnosis of schistosomiasis-related liver fibrosis have focused on difficult-to-acquire and subjective ultrasound images dependent on extensive expertise. Here we present SchistoTrackVideoNet, the first deep learning-based video model trained on easy-to-acquire standardised ultrasound video sweeps for classification of schistosomiasis-related liver fibrosis. This video-based classification model was trained and evaluated on video sweeps from 2140 participants aged 5–87 years from three districts in rural Uganda. We tested the model at a clinically-relevant sensitivity threshold (≥90%) and achieved positive predictive values of 0.0968–0.5556 for diverse presentations of liver fibrosis. Our findings show potential for the use of easy-to-acquire video sweeps for diagnosis of schistosomiasis-related liver fibrosis and our model provides a proof-of-concept for deep learning applied to liver ultrasound video for diagnosis of schistosomiasis-related liver morbidity.

## INTRODUCTION

Schistosomiasis is a neglected tropical disease prevalent in rural areas of sub-Saharan Africa (SSA), where *Schistosoma mansoni* is common. Chronic *S. mansoni* infection can lead to a complex form of liver fibrosis found in the periportal space. If left undiagnosed and untreated, severe schistosomal liver fibrosis can lead to portal hypertension and gastroesophageal varices with high mortality rates [1]. Diagnosis of schistosomal liver fibrosis is challenging and reliant on ultrasound imaging. The periportal space is a difficult area to image due to complex, overlapping anatomical structures [2]. Moreover, the presence of liver injury caused by, for example, hepatitis B or fatty liver disease may obscure clear patterns in disease presentation [3]. Diagnosis of schistosomiasis-related liver fibrosis is further complicated by presentations in ultrasound imaging being diverse, spanning diffuse and focal lesions found in multiple liver views [4]. New diagnostic tools are needed to address both the challenges in diagnosis and to fill the gap in sonography expertise in schistosomiasis-endemic areas.

Current World Health Organization (WHO) guidance for diagnosis of schistosomiasis-related liver morbidity outlines image patterns to identify presentations of liver fibrosis using B-mode ultrasound imaging [4]. The image patterns capture diffuse and focal presentations of fibrosis, with focus on first and second order branches of the main portal vein, the main portal vein itself, and the liver parenchyma [5, 6]. Considerable exposure to example cases is necessary to acquire the image patterns and to distinguish schistosomiasis-related cases from those caused by comorbidities or artifacts. Due to the lack of integrated schistosomiasis care in primary healthcare systems, opportunities to gain such expertise are rare outside of research studies [1, 7]. Due to the coinfections and comorbidities that may be found in schistosomiasis-endemic areas, and the fact that multiple fibrosis patterns can be present simultaneously, a single ultrasound image cannot capture sufficient information for a full diagnosis. Individuals affected by schistosomiasis-related liver morbidity most often have multiple presentations of liver fibrosis spanning focal and diffuse patterns [8]. Despite these limitations, current guidance and tools do not go beyond the interpretation of single ultrasound images with singular diagnoses. There is a need to investigate the applicability of models analysing ultrasound video to address challenges related to schistosomiasis-related liver fibrosis.

Despite its non-invasive nature and relatively low cost compared to other imaging modalities, there remain barriers to using ultrasound imaging that limit its use in low- and middle-income country settings [9, 10]. When coupled with the specific challenges of diagnosing schistosomiasis-related liver morbidity, more general challenges such as lack of trained personnel, lack of access to more experienced sonographers for training support and limited access to ultrasound machines prohibit diagnosis of common liver diseases via ultrasound imaging where such diagnoses are most needed. Due to the difficulties in diagnosis and expertise required, there also exists variation between assessments of schistosomiasis morbidity that may affect potential treatment avenues [11, 12]. Deep learning-based models with simple standardised video sweeps have been proposed to help address the lack of sonographer training for fetal assessments [13–17]. However, it remains to be addressed whether similar methods work for complex liver disease diagnosis. To understand the effectiveness of such methods, there is a need to understand whether deep learning-based models on ultrasound videos can perform as well or better than deep learning-based models on ultrasound images curated by expert sonographers. Additional knowledge gaps that must be addressed for schistosomiasis, and that could be applied to other health problems in low-income settings, concern how best to use outcomes from deep learning-based models to aid clinical decision support and to train new sonographers.

Here we developed the first deep learning-based models for schistosomal periportal fibrosis classification using ultrasound video. We designed a protocol comprising three standardised, easy-to-collect, liver ultrasound video sweeps that represent views of the liver including a transverse view of the right lobe, a right oblique view of the main portal vein and parasternal cross-sections of the left liver lobe. Ultrasound videos were collected as part of the community-based SchistoTrack cohort study [18, 19]. We trained and evaluated deep learning models on data from 2140 participants aged 5–87 years from 52 rural villages in Pakwach, Buliisa, and Mayuge districts of Uganda. Our proposed model was trained with a multilabel approach, using TimeSformer encoders and k-nearest neighbours (k-NN) classifiers, to make predictions of multiple presentations of schistosomiasis-related liver fibrosis simultaneously on a participant level. The aim of this work was to compare models developed on easy-to-acquire ultrasound video sweeps to models developed on (single) ultrasound images of expert-curated views of liver fibrosis patterns. We focused model evaluation on clinically-relevant metrics for diagnostics and the information needed for training and clinical decision support for sonographers in low-income settings.

## RESULTS

### Prevalence of schistosomiasis-related liver fibrosis

The total number of participants used for this analysis was 2140. The mean age of these participants was 25.3 years (standard deviation (s. d.) 18.3), and the percentage of female participants was 54.7% (1170/2140). 46.2% (988/2140) of participants were from Pakwach district of Uganda, 29.9% (640/2140) were from Buliisa district, and 23.9% (512/2140) were from Mayuge district. Table 1 shows the distribution of fibrosis patterns among the participants used for this analysis. Fibrosis patterns are assigned letters B–F, with B patterns representing diffuse fibrosis not necessarily linked to schistosomiasis; C1 and C2 patterns representing sagittal and transverse cross-sections of fibrosis in second-order branches of the main portal vein; the D pattern presenting as an echogenic ruff around the main portal vein; and E/F patterns representing severe fibrosis presenting as echogenic patches or a bird’s claw pattern stretching to the edge of the liver [4]. Since participants could have multiple patterns, the prevalence of the different combinations of patterns is summarised in Supplementary Table S1. The most prevalent pattern combinations were the B pattern alone followed by C1 and C2 together, which represented different views of the same type of fibrosis. Supplementary Figure S5 shows the distribution of the number of patterns per participant. Participants had a median of 2 (interquartile range (IQR) 1–3) patterns.

**Table 1:** Prevalence of schistosomal periportal fibrosis patterns in 2023, among the 2140 participants studied. Patterns are not mutually exclusive, therefore there are more total patterns than participants.

| Field scan pattern | Train set | Validation set | Test set | Total number of participants | Test set (2024) |
| --- | --- | --- | --- | --- | --- |
| Healthy | 75.8% (1134/1496) | 70.6% (243/344) | 81.3% (244/300) | 75.7% (1621/2140) | 0% (0/58) |
| B | 11.7% (175/1496) | 12.2% (42/344) | 11.0% (33/300) | 11.7% (250/2140) | 51.7% (30/58) |
| C1 | 13.1% (196/1496) | 18.0% (62/344) | 10.0% (30/300) | 13.5% (288/2140) | 53.5% (31/58) |
| C2 | 12.3% (185/1496) | 18.6% (64/344) | 10.0% (30/300) | 13.0% (279/2140) | 55.2% (32/58) |
| D | 5.2% (78/1496) | 7.0% (24/344) | 3.7% (11/300) | 5.3% (113/2140) | 15.5% (9/58) |
| E/F | 1.5% (22/1496) | 2.3% (8/344) | 2.0% (6/300) | 1.7% (36/2140) | 6.9% (4/58) |
| **Total participants** | 1496 | 344 | 300 | 2140 | 58 |
| **Total patterns** | 1790 | 443 | 353 | 2587 | 106 |

The ultrasound data used to train and validate the presented model was collected as part of the 2023 time point of the SchistoTrack cohort [19]. The model was tested on participant data from both the 2023 and 2024 time points. Ultrasound video data from the same participants as from the test set collected at the 2024 time point was read by a second sonographer for fibrosis patterns. The second sonographer also recorded if there were quality issues or artifacts in the ultrasound video caused by gas, fat, bone, breathing/movement, dehydration or calcification; and rated their confidence to make a clinical decision based on the video.

### Modelling pipeline

The ultrasound video sweep protocol comprised three five second long (100 frame) videos which targeted relevant anatomy for capturing periportal fibrosis liver patterns. Each acquired sweep started and ended with predefined anatomical anchors, given in Supplementary Text S.1, which did not require knowledge of schistosomiasis morbidity. Sweep one targeted views of the liver parenchyma which may capture the B, C2 and E/F patterns of fibrosis; sweep two targeted views of the main portal vein which may capture the D pattern; and sweep three targeted views of the left liver lobe which may capture the C1 pattern.

Our liver fibrosis pattern classifier, SchistoTrackVideoNet, shown schematically in Figure 1, comprised five sub-models for identification of each fibrosis pattern combined with a sixth multilabel k-NN classification model that took into account all patterns simultaneously. Each sub-model consisted of a TimeSformer encoder which takes in video clips from the relevant sweep for the target pattern, followed by two k-NN classifiers. SchistoTrackVideoNet output probabilities for each liver fibrosis pattern described in the Niamey protocol (labelled as B, C1, C2, D and E/F), given by logistic regression models that took into account probabilities from the individual sub-models and the multilabel k-NN classification model.

**Fig. 1:**
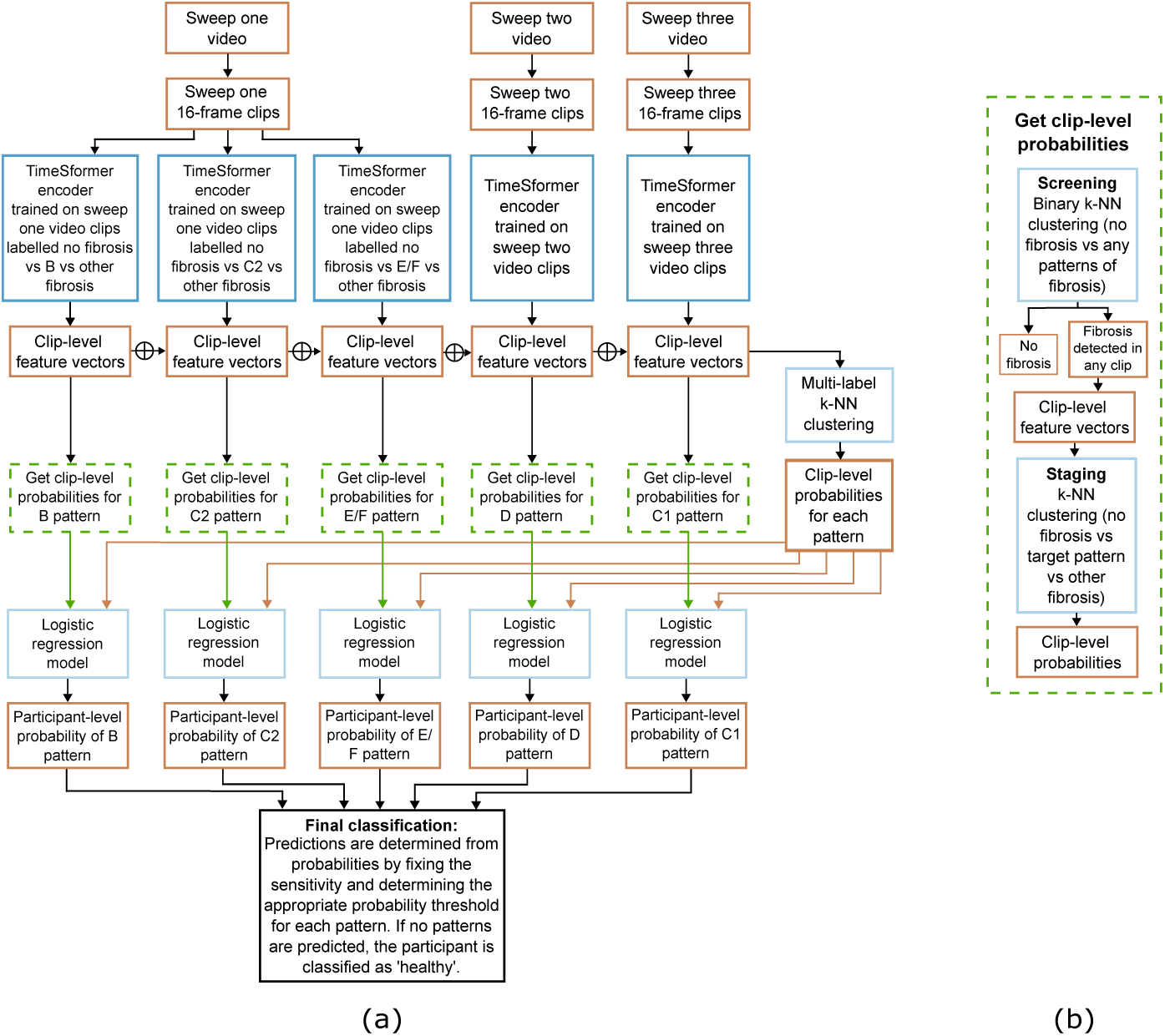
Modelling pipeline. (a) Full modelling pipeline. (b) Method to gain clip-level probabilities for each fibrosis pattern.

### Deep learning classification of liver fibrosis patterns

SchistoTrackVideoNet produced high area under the precision-recall curve (AUC-PR) values for each fibrosis pattern, particularly when taking into account the low prevalence of the patterns (Table 2). The AUC-PR values ranged from 0.3285–0.7912, which were 5.84–21.5 times higher than the prevalence of the liver patterns. Precision-recall curves are shown in Supplementary Figure S9. The area under the receiver operating curve (AUROC) values were higher than the AUC-PR values, ranging 0.9141–0.9867 among the liver fibrosis patterns. At maximum F1-score, the sensitivity for the D pattern was 0.9091, however the sensitivity was lower in the other patterns. The F1-score ranged 0.5000–0.8333.

**Table 2:** Performance of the proposed model setup for identification of each liver fibrosis pattern.

| Overall |  |  |  |  |  |  |
| --- | --- | --- | --- | --- | --- | --- |
|  | B | C1 | C2 | D | E/F |  |
| AUROC | 0.9156 | 0.9314 | 0.9141 | 0.9867 | 0.9369 |  |
| AUC-PR | 0.6448 | 0.7529 | 0.6893 | 0.7912 | 0.3285 |  |
| Prevalence | 0.1104 | 0.1003 | 0.1003 | 0.0368 | 0.0201 |  |
| AUC-PR:prevalence | 5.842 | 7.5034 | 6.8699 | 21.5072 | 16.3719 |  |
| At maximum F1-score |  |  |  |  |  |  |
|  | B | C1 | C2 | D | E/F | Healthy |
| Accuracy | 0.9197 | 0.9431 | 0.9298 | 0.9866 | 0.9799 | 0.9030 |
| F1-score | 0.6571 | 0.7119 | 0.6557 | 0.8333 | 0.5000 | 0.9405 |
| Sensitivity | 0.6970 | 0.7000 | 0.6667 | 0.9091 | 0.5000 | 0.9424 |
| Specificity | 0.9474 | 0.9703 | 0.9591 | 0.9896 | 0.9898 | 0.7321 |
| PPV | 0.6216 | 0.7241 | 0.6452 | 0.7692 | 0.5000 | 0.9385 |
| NPV | 0.9618 | 0.9667 | 0.9627 | 0.9965 | 0.9898 | 0.7455 |
| At 90% sensitivity threshold |  |  |  |  |  |  |
|  | B | C1 | C2 | D | E/F | Healthy |
| Accuracy | 0.7659 | 0.8829 | 0.7793 | 0.9699 | 0.8127 | 0.7492 |
| F1-score | 0.4615 | 0.6067 | 0.4500 | 0.6897 | 0.1765 | 0.8201 |
| Sensitivity | 0.9091 | 0.9000 | 0.9000 | 0.9091 | 1.0000 | 0.7037 |
| Specificity | 0.7481 | 0.8810 | 0.7658 | 0.9722 | 0.8089 | 0.9464 |
| PPV | 0.3093 | 0.4576 | 0.3000 | 0.5556 | 0.0968 | 0.9828 |
| NPV | 0.9851 | 0.9875 | 0.9856 | 0.9964 | 1.0000 | 0.4240 |
| At 80% sensitivity threshold |  |  |  |  |  |  |
|  | B | C1 | C2 | D | E/F | Healthy |
| Accuracy | 0.8127 | 0.8829 | 0.8629 | 0.9833 | 0.9064 | 0.8361 |
| F1-score | 0.4909 | 0.5783 | 0.5393 | 0.7826 | 0.2632 | 0.8918 |
| Sensitivity | 0.8182 | 0.8000 | 0.8000 | 0.8182 | 0.8333 | 0.8313 |
| Specificity | 0.8120 | 0.8922 | 0.8699 | 0.9896 | 0.9078 | 0.8571 |
| PPV | 0.3506 | 0.4528 | 0.4068 | 0.7500 | 0.1562 | 0.9619 |
| NPV | 0.9730 | 0.9756 | 0.9750 | 0.9930 | 0.9963 | 0.5393 |
| Image-based model |  |  |  |  |  |  |
|  | B | C1 | C2 | D | E/F | Healthy |
| Sensitivity | 0.7460 | 0.8636 | 0.8447 | 0.9143 | 0.5789 | 0.9250 |
| Specificity | 0.9544 | 0.9628 | 0.8970 | 0.9899 | 0.9831 | 0.9847 |
| PPV | 0.8704 | 0.8879 | 0.7190 | 0.8889 | 0.6111 | 0.8605 |
| NPV | 0.9015 | 0.9540 | 0.9487 | 0.9924 | 0.9807 | 0.9923 |

When the sensitivity for each liver pattern was set to 90%, sensitivity for the healthy class was 70.37% and the specificity for fibrosis patterns ranged 0.7481–0.9722, with the D pattern gaining the highest specificity. When the sensitivity for each liver pattern was set to 80%, the sensitivity for the healthy class (83.1%) and the specificity for each of the fibrosis patterns (ranging 0.8120–0.9896) was higher than when set to 90%. At 90% sensitivity, the positive predictive values (PPVs) for the fibrosis patterns ranged 0.0968–0.5556. The highest PPV was achieved by the D pattern, and the lowest was achieved by the E/F pattern, which had very low prevalence. Negative predictive values (NPVs) were ≥0.98 among all fibrosis patterns, but the NPV for the healthy class was 0.424.

When looking at t-distributed stochastic neighbour embedding (t-SNE) projections of the feature vectors given by the TimeSformer encoders used within SchistoTrackVideoNet, colouring the projections by fibrosis pattern showed some clustering by fibrosis pattern (Supplementary Figure S7). Colouring the projections by the sonographer that collected the videos (Supplementary Figure S8) showed that videos from one sonographer were clustered together but the other three were not distinct.

### Participant-level predictions of multiple fibrosis patterns

SchistoTrackVideoNet predicted multiple liver fibrosis patterns per participant. The distribution of the combinations of liver patterns that were predicted by SchistoTrackVideoNet when maximum F1-score was used is shown in Supplementary Table S3, and when the sensitivity was set to 90% the distribution is shown in Supplementary Table S4. Comparing the predicted distributions to the true distribution of the grade combinations in the test set (Supplementary Table S2), the predicted distributions more closely matched when the model was evaluated at the maximum F1-score, where the four most common pattern combinations were the same. The B pattern alone was the most predicted and the most prevalent pattern combination in all cases. When the 90% sensitivity threshold was imposed, the combinations that contained the E/F pattern were predicted more often than they appeared in the data. When comparing participants with one, two or three patterns (Supplementary Tables S5 and S6), the D pattern was missed more often when there were three patterns present, whereas the model was more sensitive to the B, C1 and C2 patterns when there were three patterns present.

### Video clips flagged as having liver fibrosis

Videos were partitioned into clips before being input to the TimeSformer encoders, and probabilities of the presence of patterns were given on a clip level before being combined by participant. For sweep one, the median probability of a B pattern peaked at clip four (Figure 2(a)), where a longitudinal section of second order portal vein branches is in view. The median probability of a C2 pattern was consistent throughout clips three, four and five (Figure 2(b), also representing the longitudinal section of second order portal vein branches where the pattern is found. For the E/F patterns, the only clips with probability distributions with non-zero upper quartiles were clips one and two (Figure 2(c)), where the gall bladder is in view. For sweep two, the peak median probability of a D pattern was in clips two and five, representing a view of the main portal vein and a view of the three segmental branches of the main portal vein (Figure 2(d)). For sweep three, the median probability of a C1 pattern stayed constant as the sweep moved through the left liver lobe (Figure 2(e)).

**Fig. 2:**
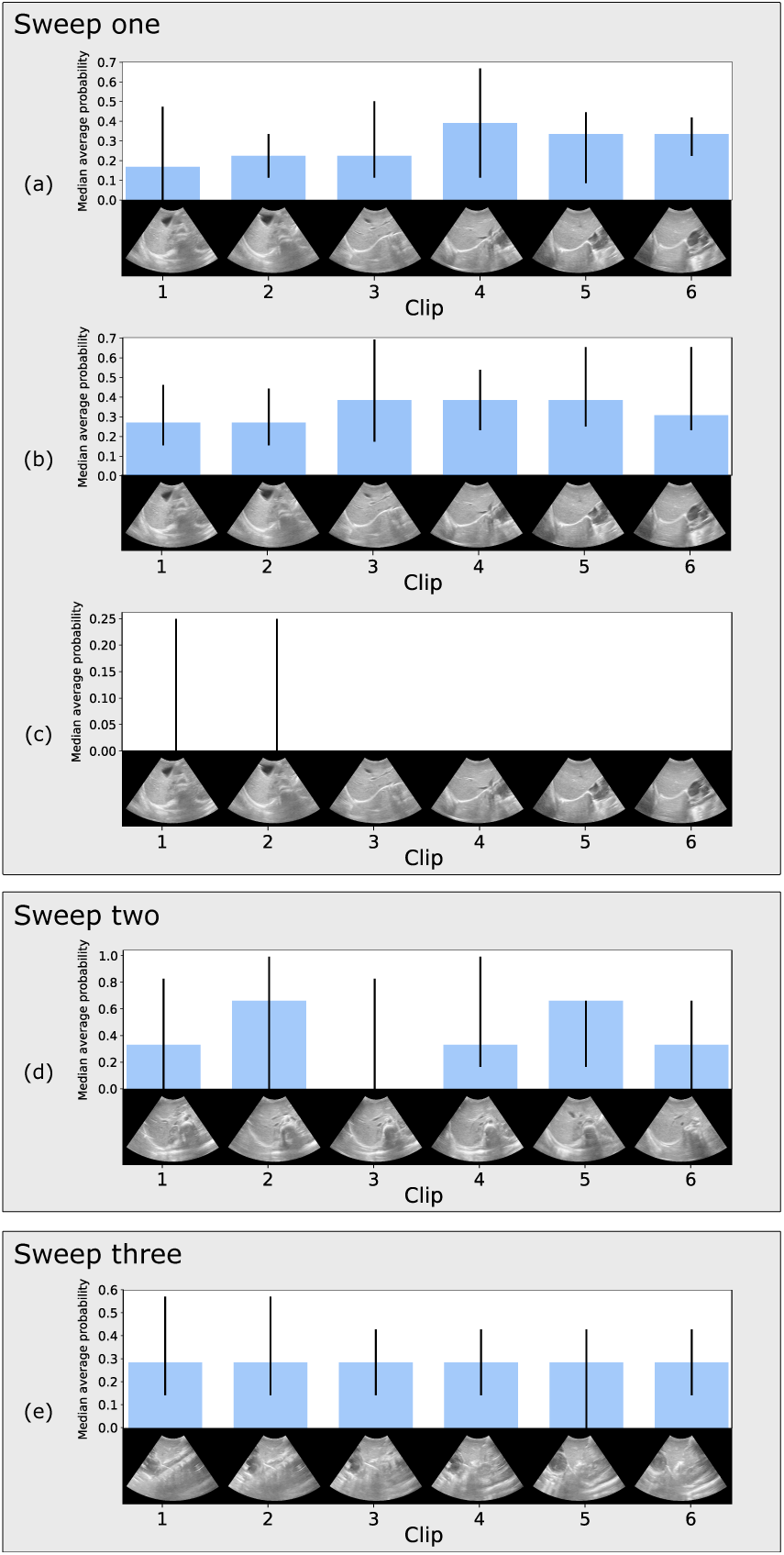
Distribution of median probability based on clip number. Each video sweep, which was 5 seconds (100 frames) long was partitioned into 16-frame clips before being input to the encoders. Error bars show the interquartile range among the probabilities for each clip number. Only videos of true cases are included. Starting frames for each video clip are shown. (a) B (with starting frames for sweep one video clips, starting with a good view of the gall bladder and ending in the cephalic position). (b) C2 (with starting frames for sweep one video clips, starting with a good view of the gall bladder and ending in the cephalic position). (c) E/F (with starting frames for sweep one video clips, starting with a good view of the gall bladder and ending in the cephalic position). The median probability was zero for all video clips, and the upper quartile was non-zero only for the first two clips. (d) D (with starting frames for sweep two video clips, starting with a good view of the main portal vein and ending with the three segmental portal branches). (e) C1 (with starting frames for sweep three video clips, which moves through the left lobe of the liver).

### Comparison with image-based model

A convolutional neural network-based image model was trained and tested on ultrasound images of curated views of fibrosis patterns, which required specific expertise in schistosomiasis morbidity to collect. At maximum F1-score, there were only small differences in sensitivity between the image-based model and SchistoTrackVideoNet, particularly in the more severe patterns (D and E/F) where the differences were 0.0052–0.0789 (Table 2). The specificity was very similar between the image-based model and SchistoTrackVideoNet evaluated at maximum F1-score, aside from for the E/F class. When the sensitivity threshold was fixed to 90%, SchistoTrackVideoNet attained higher sensitivity across the fibrosis patterns, apart from the D pattern, than the image model.

### Effect of age, sex and comorbidities

The number of missed patterns was similar between participants of different ages, sexes and with other sonography findings, apart from examples of fatty liver in the model evaluated at maximum F1-score (Table 3). There were more errors in adults than there were in children. For SchistoTrackVideoNet evaluated at 90% sensitivity, there were fewer errors in participants that did not fast and were pregnant than in the general population. Notably, adults and children had different distributions of fibrosis patterns, with adults having a higher prevalence of all patterns apart from the B pattern. There were no children in the test set that had D or E/F patterns. The sensitivity for B patterns was higher in children than in adults for both SchistoTrackVideoNet evaluated at maximum F1-score and 90% sensitivity (differences of 0.38 and 0.19 respectively). However, the sensitivity for C1 and C2 patterns was higher in adults than in children. When evaluated at a 90% sensitivity threshold, SchistoTrackVideoNet performs equally well between male and female participants (Supplementary Table S10). Slightly higher sensitivities were attained by videos from male participants than female participants (differences of 0.10–0.11), for all patterns apart from the D and E/F patterns (differences of 0–0.11). However, when evaluated at maximum F1-score, the sensitivity for all patterns was much higher in male participants than female participants (differences of 0.13–0.60), apart from the D pattern (difference of 0.11).

**Table 3:**
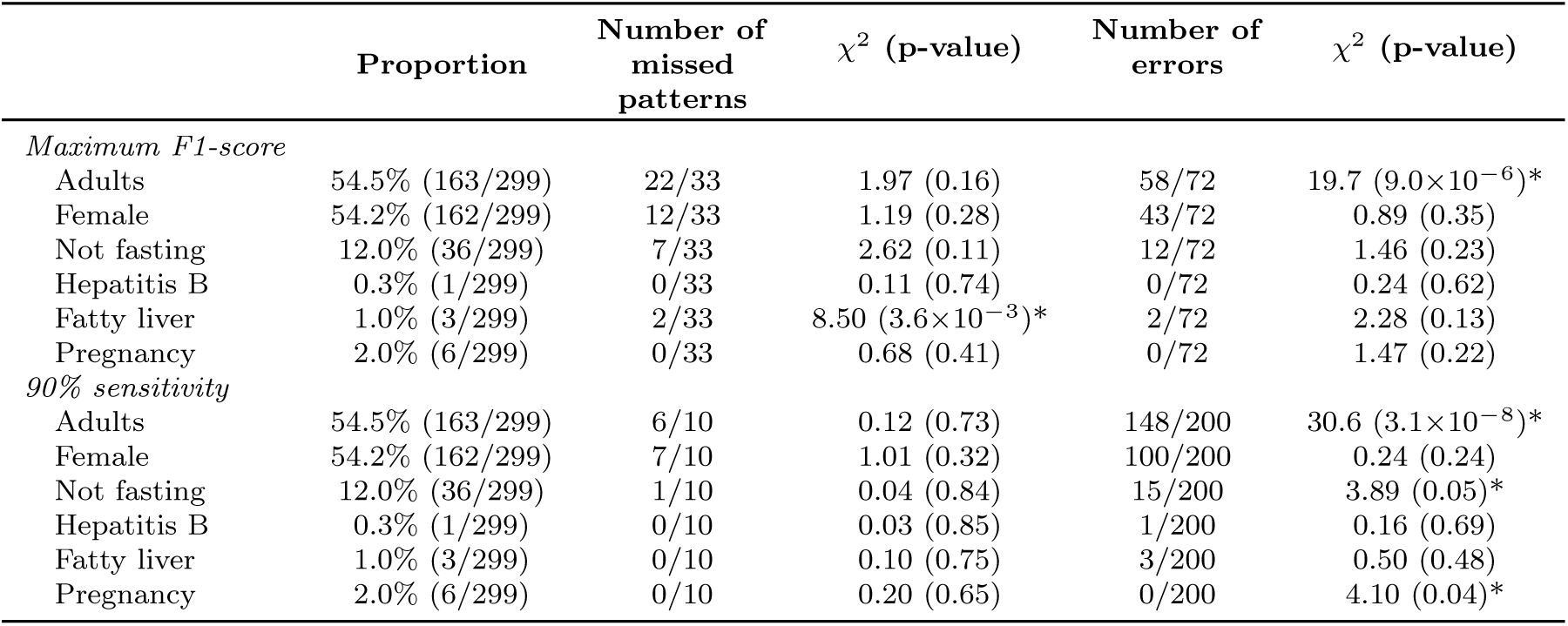
Effect of patient-level factors on model performance on the test set. A summary of the numbers of errors in adults (defined as participants aged ≥ 15 years, female participants, participants with the most common sonography findings and participants who did not fast for at least 2 hours before the ultrasound examination, for the test set.

### Model performance versus inter-sonographer agreement and effect of artifacts and clinical confidence

Ultrasound videos from 58 participants from the test set (58/300, 19.3%) were read for fibrosis patterns a second sonographer, who also recorded artifacts caused by gas, fat, bone, breathing/movement, dehydration or calcification, and scored the videos for confidence in using the videos for clinical decision-making, at the 2024 time point. For the second sonographer readings, the sensitivity for assigning each pattern at the second reading was low for all of the patterns, with the highest sensitivity being 0.4167, achieved for the B liver fibrosis pattern (Table 4). No cases of D or E/F patterns assigned in the field were also identified at the second reading from ultrasound video. SchistoTrackVideoNet evaluated at maximum F1-score gave higher sensitivity for all patterns than the sonographers at the second reading. The specificity was higher for SchistoTrackVideoNet for B and C2 patterns and higher for the sonographers for all other patterns. Compared with the same model evaluated on 2023 data (shown in Table 2), the sensitivity for SchistoTrackVideoNet evaluated at maximum F1-score on 2024 data was higher in the C1, C2 and E/F patterns. For SchistoTrackVideoNet evaluated at 90% sensitivity, the specificity was lower for 2024 data (range 0–0.51) than 2023 data (range 0.75–0.97). The PPV had a wider range for 2024 data than 2023 data, ranging 0.07–0.73.

**Table 4:**
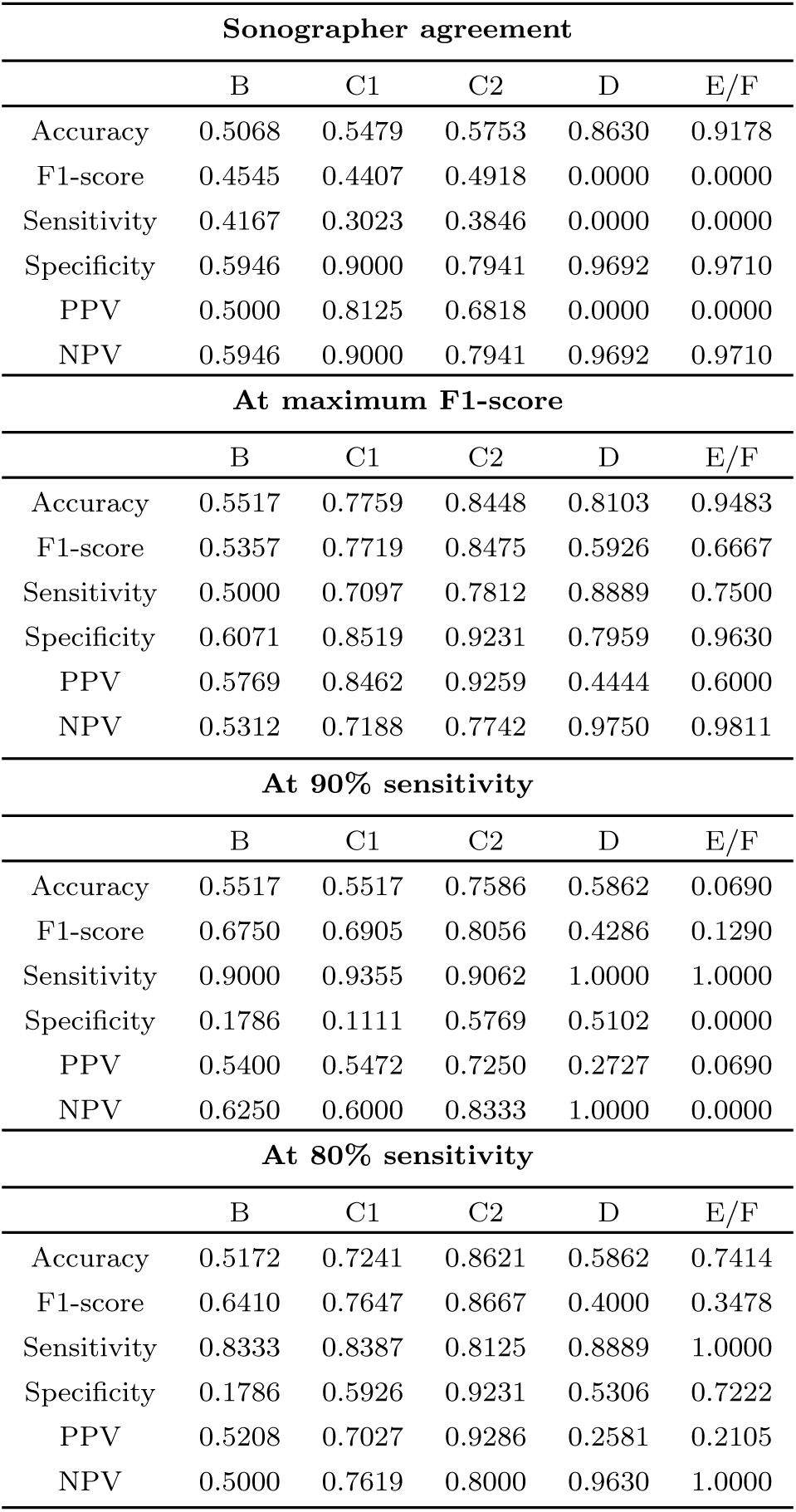
Quantitative metrics compared to sonographer agreement.

| Sonographer agreement |  |  |  |  |  |
| --- | --- | --- | --- | --- | --- |
|  | B | C1 | C2 | D | E/F |
| Accuracy | 0.5068 | 0.5479 | 0.5753 | 0.8630 | 0.9178 |
| F1-score | 0.4545 | 0.4407 | 0.4918 | 0.0000 | 0.0000 |
| Sensitivity | 0.4167 | 0.3023 | 0.3846 | 0.0000 | 0.0000 |
| Specificity | 0.5946 | 0.9000 | 0.7941 | 0.9692 | 0.9710 |
| PPV | 0.5000 | 0.8125 | 0.6818 | 0.0000 | 0.0000 |
| NPV | 0.5946 | 0.9000 | 0.7941 | 0.9692 | 0.9710 |
| At maximum F1-score |  |  |  |  |  |
|  | B | C1 | C2 | D | E/F |
| Accuracy | 0.5517 | 0.7759 | 0.8448 | 0.8103 | 0.9483 |
| F1-score | 0.5357 | 0.7719 | 0.8475 | 0.5926 | 0.6667 |
| Sensitivity | 0.5000 | 0.7097 | 0.7812 | 0.8889 | 0.7500 |
| Specificity | 0.6071 | 0.8519 | 0.9231 | 0.7959 | 0.9630 |
| PPV | 0.5769 | 0.8462 | 0.9259 | 0.4444 | 0.6000 |
| NPV | 0.5312 | 0.7188 | 0.7742 | 0.9750 | 0.9811 |
| At 90% sensitivity |  |  |  |  |  |
|  | B | C1 | C2 | D | E/F |
| Accuracy | 0.5517 | 0.5517 | 0.7586 | 0.5862 | 0.0690 |
| F1-score | 0.6750 | 0.6905 | 0.8056 | 0.4286 | 0.1290 |
| Sensitivity | 0.9000 | 0.9355 | 0.9062 | 1.0000 | 1.0000 |
| Specificity | 0.1786 | 0.1111 | 0.5769 | 0.5102 | 0.0000 |
| PPV | 0.5400 | 0.5472 | 0.7250 | 0.2727 | 0.0690 |
| NPV | 0.6250 | 0.6000 | 0.8333 | 1.0000 | 0.0000 |
| At 80% sensitivity |  |  |  |  |  |
|  | B | C1 | C2 | D | E/F |
| Accuracy | 0.5172 | 0.7241 | 0.8621 | 0.5862 | 0.7414 |
| F1-score | 0.6410 | 0.7647 | 0.8667 | 0.4000 | 0.3478 |
| Sensitivity | 0.8333 | 0.8387 | 0.8125 | 0.8889 | 1.0000 |
| Specificity | 0.1786 | 0.5926 | 0.9231 | 0.5306 | 0.7222 |
| PPV | 0.5208 | 0.7027 | 0.9286 | 0.2581 | 0.2105 |
| NPV | 0.5000 | 0.7619 | 0.8000 | 0.9630 | 1.0000 |

Very few videos among the 58 in the 2024 test set that had been read by a second sonographer had artifacts caused by gas (17.2%, 10/58), breathing/movement (6.9%, 4/58), dehydration (3.4%, 2/58) and fat (1.7%, 1/58). No videos had artifacts caused by bone or calcification. The sonographers had high confidence in making a clinical decision, such as future referrals, based on most videos (55.2%, 32/58). 32.8% (19/58) videos gave moderate confidence, and 12.0% (7/58) videos gave low confidence for this purpose. Very few videos that had missed fibrosis patterns had quality issues or artifacts, low clinical confidence, or other ultrasound findings (Supplementary Table S12).

### Sensitivity analyses

The probabilities between the clips of the video were aggregated. Two alternatives to taking the median average probability among the clip-level probabilities were considered: using the maximum probability or the mean average probability. Using the maximum and mean probabilities led to lower AUC-PR values in all patterns aside from the E/F pattern (Supplementary Table S13). For the model evaluated at maximum F1-score, the sensitivity for all patterns was highest in the model using clip-level probabilities. For the model evaluated at 90% sensitivity, the PPV was higher for all patterns when clip-level probabilities were used as opposed to the maximum or mean.

SchistoTrackVideoNet was found to generalise well to data from different districts of Uganda and performs well across a variety of low to high prevalence settings. A model with the same setup was trained on data from two districts, Pakwach and Buliisa in the west of Uganda with the highest prevalence of schistosomal liver fibrosis. The model was evaluated on data from Mayuge, which is a lower prevalence district in the east of the country. At maximum F1-score, the range of sensitivity was 0.4–0.79, compared to 0.5–0.91 when the model was evaluated on data from districts that had been seen already by the model (Supplementary Table S14). At 90% sensitivity, the range of PPV of the district model was 0.07–0.32 compared to 0.10–0.56 in the model trained and evaluated on all districts.

## DISCUSSION

Direct morbidity surveillance is an important part of achieving the WHO target for elimination of schistosomiasis as a public health problem, particularly for *S. mansoni* morbidity where infection intensity serves as a poor proxy for severity of morbidity [20–23]. Despite this, guidance for ultrasound imaging of periportal fibrosis (PPF) (the most used diagnostic method for PPF [5]) focuses on difficult-to-acquire ultrasound images [4], and training for sonographers to use this guidance is limited to research studies. We developed a deep learning-based model for classification of schistosomal PPF patterns from ultrasound video. Easy-to-acquire ultrasound video sweeps were collected using a predefined and field-validated protocol focusing on anatomical anchors. This protocol required no schistosomiasis-specific experience. Our models were trained and evaluated on 2140 participants aged 5–87 years in three districts of rural Uganda with diverse participant characteristics. We presented SchistoTrackVideoNet, a deep learning-based video model that predicts multiple patterns of schistosomal liver fibrosis with high sensitivity within an individual, high PPV, and has shown robustness to ultrasound quality issues.

SchistoTrackVideoNet performs well based on clinically-relevant quantitative metrics. At 90% sensitivity for each fibrosis pattern, PPVs of (0.0968–0.5556) were achieved. Our model results are consistent among diverse population demographics, such as across male and female participants and individuals with comorbidities. There was some increase in the number of errors in adults compared with children, though this difference may be explained by the difference in prevalence of liver fibrosis, which was more prevalent in adults. Across districts, we found that when evaluating the model setup that was trained on data from two high liver fibrosis prevalence districts (Pakwach and Buliisa) and evaluated on a third lower liver fibrosis prevalence district (Mayuge), results were largely similar to when the model had seen data from all three districts. For our models to be implemented into clinical practice, further work is needed to collect additional ultrasound video data of severe (E/F) cases that have low prevalence in a community-based setting, but may be highly prevalent in a clinical setting.

SchistoTrackVideoNet combined with the video sweep protocol was able to identify multiple patterns per participant, matching the distribution of combinations of patterns in the population when the model was evaluated at maximum F1-score. Schistosomiasis-related morbidity is complex and infection often co-occurs with other infections that affect the liver such as hepatitis B [24]. Patterns that may have causes that are not schistosomiasis (in particular the B patterns) need to be identified alongside C–F patterns since they may require different treatment strategies and are of relevance for integrated or additional surveillance programmes. PPF patterns (C–F) can co-occur within patients, and patterns are not necessarily progressive, requiring diagnostics to take into account multiple patterns at once so that diagnoses represent the biological reality in a patient. Collecting all patterns in a participant could allow future studies to investigate associations to be made between combinations of patterns and identify progression to severe outcomes such as gastroesophageal varices.

Our ultrasound video sweep procedure coupled with SchistoTrackVideoNet could enable large-scale collection of ultrasound data for schistosomiasis-related morbidity surveillance. We evaluated our model at 80% sensitivity to match the minimum recommended sensitivity in the target product profile for schistosomiasis diagnostics for monitoring and evaluation of schistosomiasis control programmes [25], yielding strong sensitivity particularly in the D and E/F patterns. The ultrasound video sweep protocol captures the views of the liver that are required for diagnosing schistosomiasis-related morbidity, where sweep one targets the liver parenchyma in the transverse view, sweep two targets the main portal vein in an oblique view and sweep three targets the left lobe of the liver in the sagittal view. For sonographers, agreement between diagnoses based on second readings of ultrasound videos was low compared to second readings of ultrasound images, which has been shown to give *κ* values between moderate to almost perfect agreement [11, 26, 27]. However, there is far less disparity, if any, between the performance from models on ultrasound images versus videos, despite the images being targeted to specific fibrosis patterns in curated views of the liver. One possible explanation could be that deep learning models designed for video may capture long-range connections between frames and evaluate the quantity of information in a video, however sonographers are not trained to recognise fibrosis patterns from video or may not have the capacity to quickly process so much information in detail. In the under-resourced settings in which schistosomiasis is endemic, there are very few sonographers that have expertise in diagnosing schistosomiasis-related liver morbidity. Given that the ultrasound video sweep procedure outlined in this paper relies only upon basic knowledge of liver anatomy sonography to identify the anatomical landmarks that define the start and end of each sweep, rather than schistosomiasis-specific expertise, our procedures could potentially be used to enable non-specialist sonographers to collect ultrasound data for research studies. Currently, WHO guidance only advises ultrasound images to be collected for assessment of schistosomiasis morbidity, but our analysis suggests that ultrasound video sweep procedures should be considered in future WHO guidance.

We comprehensively evaluated SchistoTrackVideoNet using multiple clinically-relevant metrics with the outlook of supporting clinical decision-making. Clinical decision support systems for ultrasound imaging can provide increased diagnostic skills and accuracy both in training and in the clinic [28, 29]. Criteria for clinical decision support include sufficient generalisability, automation and integration into existing clinical systems [30, 31]. Assessing generalisability between sites is important to avoid a significant drop in performance [32]. Our study has shown that the model setup, when trained on two districts (Pakwach and Buliisa), performs comparably on the third district (Mayuge) as when the model was trained on all three districts, suggesting potential generalisability between sites with different PPF prevalences. Additionally, our models have been trained on data from six sonographers on a diverse population across age and with comorbidities, providing a foundation for future evaluation in other populations. Possible use cases for these models for clinical decision support tools for ultrasound video of PPF patterns include assisting in uncertain diagnoses due to artifacts or if there is disagreement among readers. In particular, our models appear robust to the types of quality issues assessed in our study, including those caused by gas, breathing/movement, dehydration and fat. Since sonographers with less training and fewer years of experience are less likely to identify artifacts on an ultrasound scan [33], our models could be useful in assisting such sonographers. Although we have tested the model between geographical sites with differing demographics and PPF prevalences, further studies should be done to ensure cross-country generalisation. In addition, future studies should seek to validate these models as clinical decision support tools by integrating risk prediction to perform triage, or generation of reports from ultrasound video that could be used by clinicians to inform treatments or patient follow-up. Ultimately, implementation studies also will be needed to test the impact of such systems on clinical practice [34].

Key strengths of this study lie in the large, diverse dataset, presentation of a standardised protocol for ultrasound video, high performing models that were comparable to image-based classification models, and multilabel classification allowing for biologically meaningful outputs. However, our study has several limitations. There were very few examples of the E/F fibrosis patterns in the dataset, reflecting the prevalence of E/F patterns in the study districts given survivor bias induced by high mortality rates associated with such severe patterns [7]. A larger sample or sampling within clinics would be needed to address this limitation. Further studies are needed to test the generalisability of the presented models to data from other ultrasound machines, since those that are procured locally may be of lower quality. In addition to this, the generalisability of the models to other geographic settings, and to sonographers with no schistosomiasis expertise should be studied.

Here we present the first deep learning-based model for classification of liver fibrosis patterns from ultrasound video. The multilabel model presented was able to predict multiple liver fibrosis patterns from a simple ultrasound video sweep procedure and was tested at a clinically-meaningful sensitivity threshold. This work shows that standardised, easy-to-collect ultrasound video sweeps contain sufficient information for diagnosis of schistosomal liver fibrosis patterns, advocating for the inclusion of ultrasound videos and machine learning tools in revisions of WHO protocols for schistosomal liver fibrosis assessment. With further clinical validation and additional field implementation studies in other geographic settings, we anticipate that this approach will provide the basis for automated clinical decision support for schistosomal periportal fibrosis and large-scale training of sonographers in under-resourced areas.

## METHODS

### Study design

The SchistoTrack cohort is a prospective, longitudinal community-based study of individuals. The first year of observation was conducted in 2022 and there is annual follow-up planned until 2027 [18, 19]. The cohort is based in three districts of rural Uganda: Pakwach, Buliisa and Mayuge. Two of the districts, Pakwach and Buliisa, are in the west of Uganda, bordering the White Nile and Lake Albert respectively. Buliisa and Mayuge both have hospitals in the district, however the highest level of health centre that Pakwach had was a Health Centre IV. Sonography training in Uganda does not focus on identification of schistosomiasis-related morbidity, and so this training is largely done by research studies. Therefore, in rural areas of Uganda there is a lack of sonographers trained to identify periportal fibrosis.

Households were sampled from a local register for participation in the study from 52 villages among the three districts. Uniform random sampling was used to select 40 households per village, with a further 30 households sampled to account for nonparticipation. One adult (aged 18 or older) and one child (aged younger than 18) from each household were chosen by the household head or lead wife to be a clinical participant in the SchistoTrack cohort. This study focuses on participants recruited in 2023 and 2024. Further details on the cohort can be found at [18].

### Liver pattern diagnoses

Philips C5-2 curvilinear transducers were used with the Lumify Application (version 4.0.1) connected to tablets with Android 9 Pie. The abdomen setting was used when saving images. All videos were collected in lossless digital imaging and communications in medicine (DICOM) format. Scans were performed alongside a survey programmed in Open Data Kit (version 2023.2.4) on another Android 9 Pie tablet, filled out by an assistant to the sonographer who could speak the local language and was from the district where the data collection was taking place on that day. Each participant was seen by one expert sonographer and examined within their village. Four sonographers with 5-20 years of experience on diagnosing schistosomal liver fibrosis conducted the examinations.

Diagnoses of image patterns as described in the Niamey protocol [4] were performed and ultrasound images of each pattern that was found were saved and labelled. There were three probe positions from which images of fibrosis patterns could be captured: the sagittal left parasternal view, a transverse view, and an oblique view of the portal vein. Image patterns were assigned letters from A to F, representing different presentations of schistosomal liver fibrosis (C–F) or related fibrosis (B). B patterns, which appear in the transverse view, were separated into three categories: B0 (feather streaks) which appears as lines across the liver; B1a/b (flying saucers or starry sky) which appears as solid white dots and small dots on the field of view; and B2 (spider thickening) which appears as thickening of many second order portal vein branches. C–F patterns, which represent periportal fibrosis (PPF), were in five categories: C1 (prominent peripheral rings) which appears as rings with no echogenic centre in the sagittal left parasternal view; C2 (prominent pipestems) which appears as brightness along a horizontal cross-section of a vessel in the transverse view; D (ruff) which appears as thickness around the main portal vein in the oblique view; E (patches) which appears as a blocked vessel which has no lumen, found in the transverse view; and F (bird’s claw) which appears as a blocked vessel with no lumen that is stretched to the periphery, also found in the transverse view.

### Ultrasound video sweep protocol

The ultrasound video data collection procedure was broken down into three 5-second (100-frame) sweeps, with anatomical anchors defining the start and end of each sweep. The full protocol is given in Supplementary Text S.1. Sweep one aimed to target transverse cross-sections of the liver parenchyma and views of the gall bladder; sweep two targeted views of the main portal vein and portal branches; and sweep three aimed to capture sagittal views of the left liver lobe. Since the B, C2, E and F patterns are predominantly found in the transverse view of the liver parenchyma, it was expected that these would be found in sweep one. The D pattern is defined as ruffing around the main portal vein, therefore a D pattern would be found in sweep two. Finally, the C1 pattern is found in the sagittal cross-section, therefore would be expected to be found in sweep three.

Three separate sweeps were captured rather than a single video in order to provide more anatomical anchors without needing annotation. Sonographers first undertook an unrecorded preliminary scan of the participant before capturing the video sweeps. This would allow the sonographers to assess the sizes of the organs and the presence of comorbidities, understand whether any adjustments were needed for the starting landmarks, and ultimately allow them to capture each sweep within the required 5second timeframe. Sonographers were expected to delete and repeat videos where an error had been made, or if they had been disturbed or interrupted during the recording.

### Labelling

Each participant could be assigned multiple patterns of schistosomal liver fibrosis at the point of the ultrasound examination using the sonography survey. Therefore, assigning a single label per participant could ignore fibrosis stages that could be from multiple causes, and therefore might miss important clinical outcomes that a participant could experience. Therefore, for analysis in this paper, all patterns assigned on the sonography survey were used as participant-level labels to derive a multi-label video model.

### Second readings, quality issues, clinical confidence and comorbidities

After data collection in 2024, sonographers were assigned participants at random to read their ultrasound video for liver fibrosis patterns and provide comments on video quality, artifacts and clinical confidence. Sweep one ultrasound videos were converted to MP4 format and played by the second sonographer using standard software such as VLC media player. The second sonographer was asked to record every pattern of fibrosis that appeared in that video using Google Sheets. For all videos, the second sonographer also reported on artifacts and clinical confidence. The artifact types that were given were gas, fat, breathing/movement, bone, calcification and dehydration. Gas was included as it may cause posterior enhancement; fat as it may cause excessive brightness; breathing/movement as it may cause blurring; bone as it may cause a posterior shadow; and calcification and dehydration were included as they may appear with excessive speckle. There was also space for the second sonographer to add other types of artifacts or quality issues in free text or any other comments.

The second sonographer was asked to rate their clinical confidence as low, medium or high. Low confidence was defined in the SchistoTrack standard operating procedure (SOP) as a sonographer would not use that video to take clinical actions, and this would require another scan. Moderate confidence was defined as a sonographer would use that video to take clinical actions, but more ultrasound imaging information would be helpful. High confidence was defined as a sonographer would use that video to take clinical actions and it is clear what actions were needed.

### Study participants

A total of 2140 participants were studied as part of this analysis. At the 2023 timepoint, 3219 participants attended biomedical data collection, of which 3095 had both sonography survey data and ultrasound images or videos collected. 2923 of these participants had three ultrasound video sweep videos available, and 2617 of these participants had videos that were the correct length (5 seconds). 2337 of these participants fasted for at least 2 hours before data collection. The healthy class was defined as participants who did not have any ultrasound findings (such as oesophageal varices or hepatitis B-like liver) or liver fibrosis patterns present. There was a total of 1621 participants in the healthy class, of which 766 were children (aged *<* 15) and 855 were adults (aged ≥ 15). For the liver fibrosis patterns classes there was a total of 519 participants. 185 participants with liver fibrosis patterns were children (aged *<* 15), and 334 participants with liver fibrosis patterns were adults (aged ≥ 15). A flowchart showing the participants excluded at each step is shown in Supplementary Figure S4. For participants with patterns of liver fibrosis, train, validation, and test splits were randomly assigned in a ratio of 70:20:10 until there was representation of every class in each split. The healthy participants were assigned to splits randomly in a ratio of 70:20:10.

To assess the effect of video quality, artifacts, clinical confidence and comorbidities on the model predictions, and to compare to the sonographer second readings, a subset of participants that were not used to train the model were selected from 2024 data. These participants were selected from the 2023 test set. 75.7% (259/342) participants from the 2023 test set had ultrasound video data and sonography survey data from the 2024 time-point. Only participants with liver fibrosis patterns in 2024 were considered, since these were the participants for whom second readings were available. 29.3% (76/259) participants that had ultrasound video data and sonography survey data at the 2024 time-point among the 2023 test set had liver fibrosis patterns at the 2024 time-point. 96.1% (73/76) of these participants had data on second readings, quality issues and clinical confidence in 2024.

### Data cleaning

Python 3.10 was used for all processing. The pixel array was extracted from the DICOM files produced by the Philips Lumify application using pydicom. The text surrounding the field of view was removed and the pixel value set to zero, using OpenCV (version 4.8.1.78).

### Multilabel modelling

Figure 1 shows a diagram of the multilabel deep learning-based video model used to predict liver fibrosis patterns from ultrasound video, called SchistoTrackVideoNet. Video clips from each sweep were passed through encoders to generate feature vectors. These feature vectors were then passed through binary k-NN classification ‘screening’ models that discriminated between participants with and without liver fibrosis patterns. Then, participants who had been classified as having liver fibrosis were passed on to ‘staging’ models to determine the pattern of fibrosis that was present. Each ‘staging’ model output a probability of the presence of the pattern in question per clip, based on the number of neighbours that were a member of that class. Another probability per pattern per clip was given by a sixth multilabel k-NN classification model that took into account feature vectors from all five encoders. The two vectors of probabilities were input to a logistic regression model to output a final probability for presence of that fibrosis pattern on a participant level.

The backbone encoder selected for SchistoTrackVideoNet was the TimeSformer [35], and is used to generate feature vectors from ultrasound video clips. The TimeSformer is a state-of-the-art encoder for video classification. The five TimeSformer encoders were each trained in a fully supervised manner, as described in the original TimeSformer paper [35]. The tube masking strategy, described in [36] was used. For input to the TimeSformer encoder, ultrasound videos first had to be partitioned into clips. The chosen length of these clips was 16 frames, and from these every other frame was sampled before input to the encoder.

For training of the TimeSformer encoders, a single label was allowed per video, despite each video having multiple labels. Therefore, five encoders were trained, each targeting classification of a specific liver fibrosis pattern. Since sweep one contained multiple patterns, three separate backbone encoders on sweep one video, with the encoders corresponding to B, C2 and E/F patterns respectively. For the encoder trained to classify the B fibrosis pattern, the videos were labelled with B (if there was a B fibrosis pattern present), ‘other fibrosis’ (if any other fibrosis was present), and ‘healthy’ (if there was no fibrosis present). The C2 and E/F encoders were trained using the same videos but labelled with the C2 and E/F pattern in place of the B fibrosis pattern where appropriate. In a similar manner, for the sweep two encoder, participants with a D fibrosis pattern were labelled as a D pattern, and participants with fibrosis but not a D fibrosis pattern were labelled as ‘other fibrosis’. The sweep three videos were labelled as a C1 fibrosis pattern if this pattern was present, and the ‘other fibrosis’ and healthy labels were defined as before. The ‘other fibrosis’ class acted as a dummy class to avoid grouping healthy and fibrotic participants in the same class. In addition to the sweep models, an additional multi-sweep, multilabel kNN classification was performed on concatenated feature vectors from the three sweep encoders. This was done to investigate if there was a performance improvement if the sweeps were considered together. To produce final probabilities, a logistic regression model for each pattern was trained on clip-level output probabilities from the relevant sweep encoder and the clip-level probabilities from the multi-sweep, multilabel model corresponding to that pattern. To gain classifications, decision thresholds were fixed at 80% sensitivity, 90% sensitivity, and maximum F1-score for each pattern, outputting a vector of classifications for each participant. If no patterns were identified, the participant was classified as healthy.

### Hyperparameter tuning

The batch size, initial learning rate and weight decay were tuned using the 20 iterations of Bayesian hyperparameter tuning, optimising for F1-score on the 2023 validation set. For the batch size, values of 2, 4, 8 and 16 were tried. For the initial learning rate, values between 0.0001 and 0.01 were tried. For the weight decay, values between 1 × 10^−5^ and 1 × 10^−3^ were tried. Stochastic gradient descent was used to optimise model weights.

### Performance metrics

t-distributed stochastic neighbour embedding (t-SNE) plots were created for the feature vectors given by each encoder that was trained as part of the model development. area under the receiver operating curve (AUROC), area under the precision-recall curve (AUC-PR) and AUC-PR:prevalence ratio are reported. Quantitative measures of model performance were computed at fixed decision thresholds at 80% and 90% sensitivity for each fibrosis pattern and at maximum F1-score for each pattern; these measures were accuracy, F1-score, sensitivity, specificity, positive predictive value (PPV) and negative predictive value (NPV). These metrics were reported for each liver fibrosis pattern separately. The sensitivity of the multilabel model described in this paper for each pattern was compared to the sensitivity of a convolutional neural network-based model trained and tested on ultrasound images of curated views of the liver fibrosis patterns. The error distributions among videos that had artifacts, were from participants with comorbidities or other ultrasound findings, and differing clinical confidence scores were computed and reported. Before being consolidated to a participant level, probabilities were computed on a clip level, and so each video clip from the three sweeps had predictions for fibrosis patterns. Therefore, the distributions of probabilities among the clips given to the encoders was reported, to understand whether predictions were being made when certain anatomies were present in the ultrasound video.

### Sensitivity analyses and model comparisons

To compare strategies for aggregating the clip-level probabilities for input to the logistic regression models, taking the mean and maximum clip-level probabilities was done, instead of all probabilities. To compare the model performance between age categories (adults, defined as participants aged ≥ 15 and children, defined as participants aged *<* 15) and sexes, results disaggregated by these categories are reported. Participants came from three districts of Uganda, each with differing characteristics, including mass drug administration history, ecological characteristics, and prevalence of PPF. To assess the generalisability of the proposed model setup between sites, a model was trained on the two higher prevalence districts in the west of Uganda, Pakwach and Buliisa, and tested on the lower prevalence district of Mayuge. This district split was chosen to ensure that there was enough data for each class for training the model.

## Data availability

Image and video data are protected and are not available due to data privacy laws, restrictions in ethics approvals and participant agreements associated with the ongoing SchistoTrack Cohort study. These restrictions prevent open sharing of raw images or videos outside the study and individual-level participant information that may identify individuals.

## Code availability

Code has been provided as supplementary material.

## Acknowledgments

We are thankful for the involvement of our study participants and the SchistoTrack teams, especially the sonographers, nurses and surveyors. We would also like to thank the Uganda Ministry of Health, local district leaders, focal health workers, and village health teams. We are grateful for the constant involvement through the study timepoints and community engagement meetings of our study participants. Special thanks also to the SchistoTrack and Noble Groups in Oxford for everyday discussions and feedback, and for the feedback from Bartek Papiez.

## Author contributions

Conceptualisation: EO, AN, GFC. Data curation: EO, VA, SM, TM, BNt, BNa, and GFC. Formal analysis: EO. Funding acquisition: EO and GFC. Investigation and Methodology: EO, AN, and GFC. Project administration: BNa, NBK, and GFC. Resources: GFC. Software: GFC. Supervision: AN and GFC. Validation: EO, VA, SM, TM, BNt, and GFC. Visualisation: EO. Writing - original draft: EO. Writing - review & editing: EO, VA, SM, TM, BNt, BNa, NBK, AN and GFC.

## Declaration of interests

All authors declare no competing interests.

## Ethics approvals

Data collection and use were reviewed and approved by Oxford Tropical Research Ethics Committee (OxTREC 509-21), Vector Control Division Research Ethics Committee of the Uganda Ministry of Health (VCDREC146), and Uganda National Council of Science and Technology (UNCST HS 1664ES). Written informed consent was obtained from adult participants aged 18 years and older, who also provided written consent on behalf of verbally assented children with fingerprint assent. Older children also provided written consent in addition to the adult consent on their behalf.

## Funding

E.S.O. received funding from the UKRI EPSRC as a DPhil studentship (2593890) associated with project (EP/S02428X/1). This research was funded in part by the UKRI EPSRC [EP/X021793/1]. For the purpose of Open Access, the author has applied a CC-BY public copyright licence to any Author Accepted Manuscript version arising from this submission. NDPH Pump Priming Fund, John Fell Fund, Robertson Foundation, UKRI EPSRC (EP/X021793/1) grants were awarded to G.F.C.

## Notes

### Competing Interest Statement

The authors have declared no competing interest.

