## Supplements for "SchistoTrackVideoNet: multilabel deep learning-based classification of schistosomal periportal fibrosis from ultrasound video"

### Supplementary Information S

#### S.1 Ultrasound video data collection protocol

- Before beginning, confirm that the cineloop is set to 5 seconds.
- Three separate recordings/cineloops should be taken. Each will represent one sweep.
- The purpose of these three recordings is to capture all necessary anatomy and views to identify patterns of periportal fibrosis.
- Cineloops will only start after there is a good view of target anatomy.
- You are allowed to lift the probe in between sweeps to adjust the position and find the anatomy on each participant.
- However, DO NOT lift the probe once the recording or sweep starts.
- Adjust the speed of the sweeps based on the information about liver size gathered from the superficial scan.
- DO NOT lift the probe off the participant's body during the recording.
- DO NOT stop the recording early.
- If you make a mistake, delete the cineloop with errors and start again before moving to the next cineloop.

**Sweep one**

1. Place the probe just below the sternum in the transverse position aligning with the midclavicular line.
2. Adjust the probe position until there is a good view of the gall bladder.
3. If the gall bladder is not visible at all then adjust the probe to where the gall bladder would be located approximately.
4. Start recording.
5. Perform a sweep moving the probe to the cephalic position.
6. Stop and save recording.

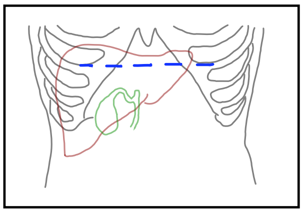

**Fig. S1**: **Starting position of sweep one.** A transverse view of the liver parenchyma. Adapted from the Niamey protocol [4].

**Sweep two**

1. Place the probe in the right oblique position (45 degrees) aiming at the main portal vein.
2. Adjust the probe so that the main portal vein is coming in at an angle and in good view.

3. Start recording.
4. From the main portal vein view, continue in oblique view and turn the probe to transverse to see the first three branches. **Always look at the branches even in healthy participants.**
5. Stop and save recording.

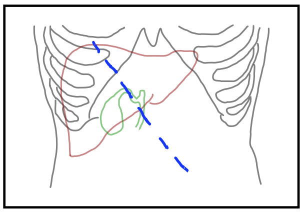

**Fig. S2**: **Starting position of sweep two.** An oblique view of the main portal vein. Adapted from the Niamey protocol [4].

##### Sweep three

1. Place the probe in the sagittal left parasternal position (90 degrees).
2. Ensure you have a view of the left lobe of the liver. Usually, this will be the whole lobe in healthy participants, but may not be the case in people with lobe enlargement.
3. Start recording.
4. Move/sweep the probe until you reach the edge of the left lobe. Make sure the sweep shows the entire left liver lobe.
5. Stop and save recording.

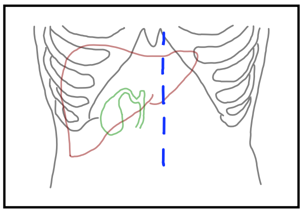

**Fig. S3**: **Starting position of sweep three.** A sagittal view of the left liver lobe. Adapted from the Niamey protocol [4].

#### S.2 Participants

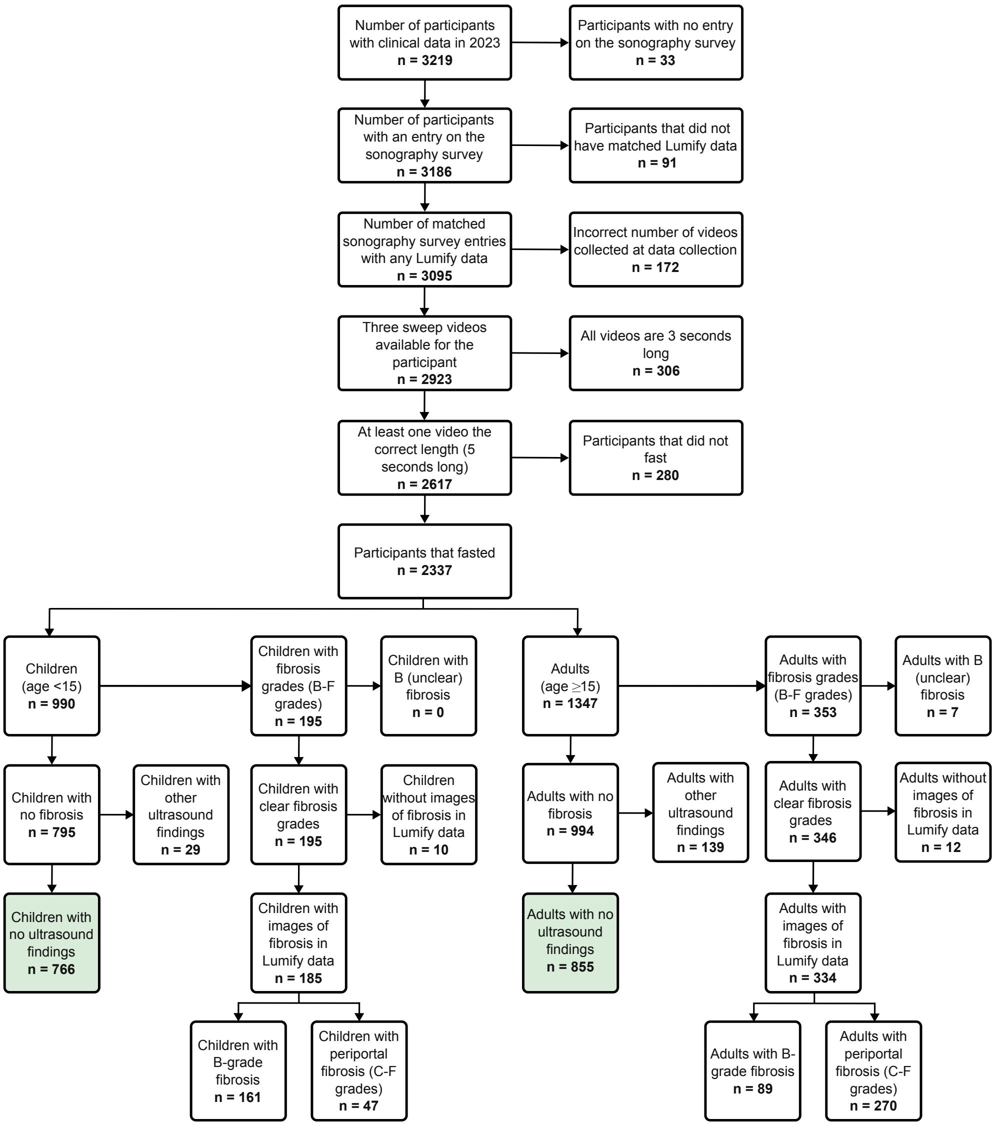


**Fig. S4**: **Participant flowchart.** Green boxes represent participants included as part of the 'no fibrosis' class. B-pattern and C–F pattern fibrosis are not mutually exclusive.

#### S.3 Combinations of patterns

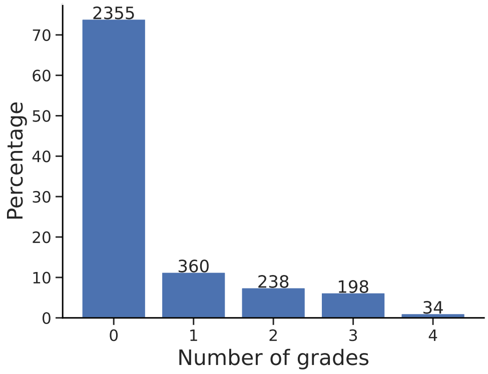


**Fig. S5**: **Distribution of number of patterns per participant.** Overall distribution on all 2023 participants.

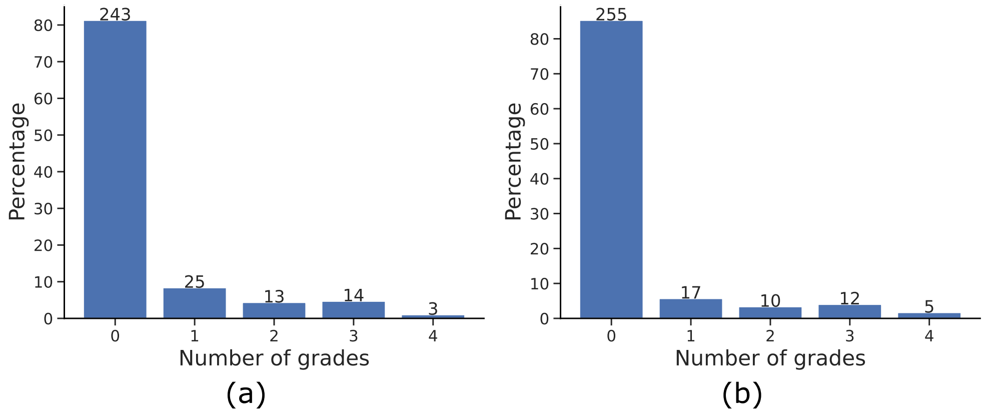


**Fig. S6**: **Distribution of number of patterns per participant.** (a) True distribution of number of patterns from the test set. (b) Predicted distribution of number of patterns from the test set, when the model is evaluated at maximum F1-score.

**Table S1**: **Counts and percentages of different fibrosis pattern combinations.** The remaining 75.4% (1621/2140) participants used in this analysis had no fibrosis patterns.

| Combination of patterns | Percentage of participants |
|---|---|
| B | 9.4% (202/2140) |
| C1, C2 | 6.4% (137/2140) |
| C1, C2, D | 2.9% (62/2140) |
| B, C1, C2 | 1.5% (33/2140) |
| C1, C2, D, E/F | 0.9% (19/2140) |
| D | 0.6% (13/2140) |
| C1, C2, E/F | 0.6% (12/2140) |
| C1 | 0.5% (11/2140) |
| C2, D | 0.3% (6/2140) |
| C1, D | 0.2% (4/2140) |
| B, C1 | 0.2% (4/2140) |
| B, C2 | 0.1% (3/2140) |
| B, C1, C2, D | 0.1% (3/2140) |
| D, E/F | 0.1% (2/2140) |
| C2 | 0.1% (2/2140) |
| B, D | 0.1% (2/2140) |
| E/F | 0.05% (1/2140) |
| B, C1, C2, E/F | 0.05% (1/2140) |
| B, C1, D | 0.05% (1/2140) |
| B, C1, C2, D, E/F | 0.05% (1/2140) |

**Table S2**: **Counts and percentages of different fibrosis pattern combinations, in the test set.** The remaining 81.3% (243/299) participants used in this analysis had no fibrosis patterns.

| Combination of patterns | Percentage of participants |
|---|---|
| B | 8.0% (24/299) |
| C1, C2 | 3.7% (11/299) |
| C1, C2, D | 2.0% (6/299) |
| B, C1, C2 | 1.7% (5/299) |
| C1, C2, E/F | 1.0% (3/299) |
| C1, C2, D, E/F | 0.3% (1/299) |
| C1, D | 0.3% (1/299) |
| D | 0.3% (1/299) |
| B, C2 | 0.3% (1/299) |
| B, C1, C2, D | 0.3% (1/299) |
| B, C1, C2, E/F | 0.3% (1/299) |
| B, C1, C2, D, E/F | 0.3% (1/299) |

**Table S3**: **Counts and percentages of different fibrosis pattern combinations, as predicted by the model evaluated at maximum F1-score for each pattern.** The remaining 81.6% (244/299) participants were predicted as having no fibrosis patterns.

| Combination of patterns | Percentage of participants |
|---|---|
| B | 7.0% (21/299) |
| C1, C2 | 2.0% (6/299) |
| C1, C2, D | 2.0% (6/299) |
| B, C1, C2 | 2.0% (6/299) |
| B, C1, C2, D | 1.3% (4/299) |
| C2 | 1.3% (4/299) |
| B, C1, C2, D, E/F | 0.7% (2/299) |
| B, C1 | 0.7% (2/299) |
| C1, C2, E/F | 0.3% (1/299) |
| C1, C2, D, E/F | 0.3% (1/299) |
| B, E/F | 0.3% (1/299) |
| B, C1, C2, E/F | 0.3% (1/299)) |

**Table S4: Counts and percentages of different fibrosis pattern combinations, as predicted by the model set at 90% sensitivity.** The remaining 58.2% (174/299) participants were predicted as having no fibrosis patterns.

| Combination of patterns | Percentage of participants |
|---|---|
| B | 9.0% (27/299) |
| B, C1, C2, E/F | 9.0% (27/299) |
| B, C2 | 4.3% (13/299) |
| B, C1, C2, D, E/F | 4.0% (12/299) |
| C2 | 4.0% (12/299) |
| B, C1, C2 | 2.3% (7/299) |
| C1, C2, D, E/F | 2.0% (6/299) |
| B, E/F | 2.0% (6/299) |
| B, C2, E/F | 1.7% (5/299) |
| C1, C2 | 1.3% (4/299) |
| C1, C2, E/F | 1.0% (3/299) |
| E/F | 0.7% (2/299) |
| C2, E/F | 0.3% (1/299) |

#### S.4 t-SNE plots

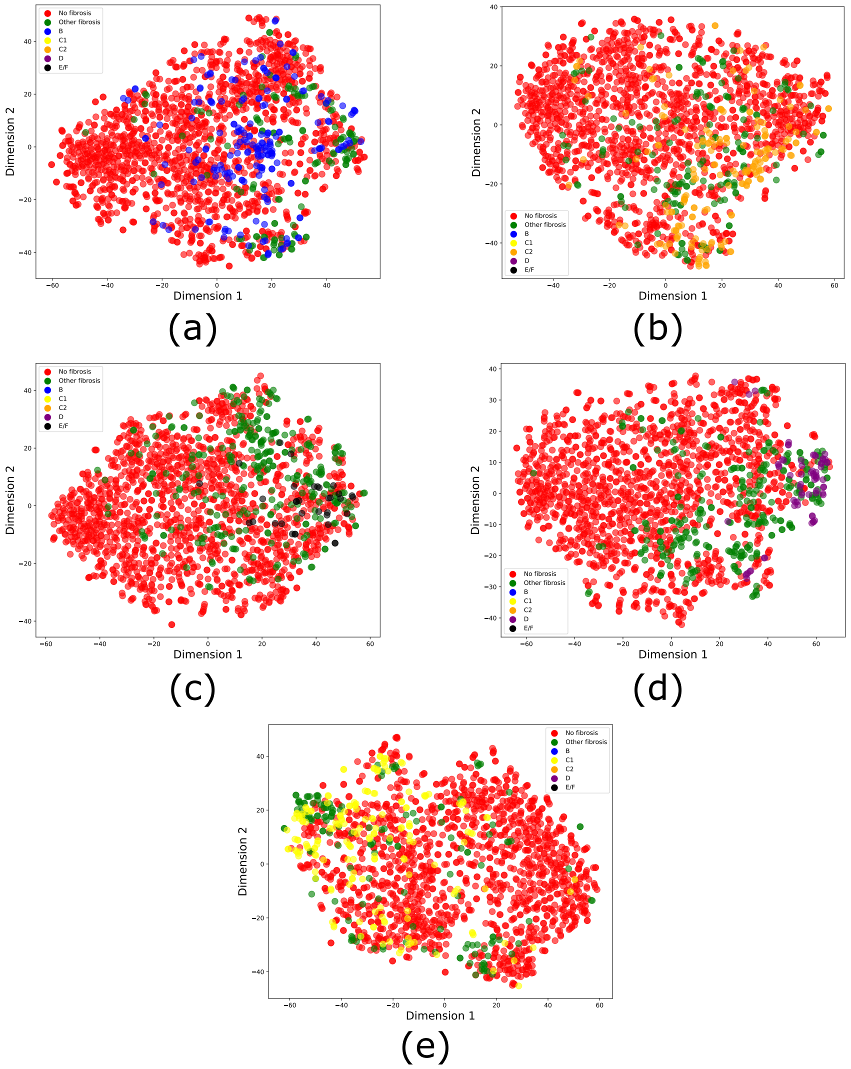


**Fig. S7**: **t-SNE coloured by fibrosis pattern.** (a) Encoder trained for B pattern. (b) Encoder trained for C2 pattern. (c) Encoder trained for E/F pattern. (d) Encoder trained on sweep two video. (e) Encoder trained on sweep three video.

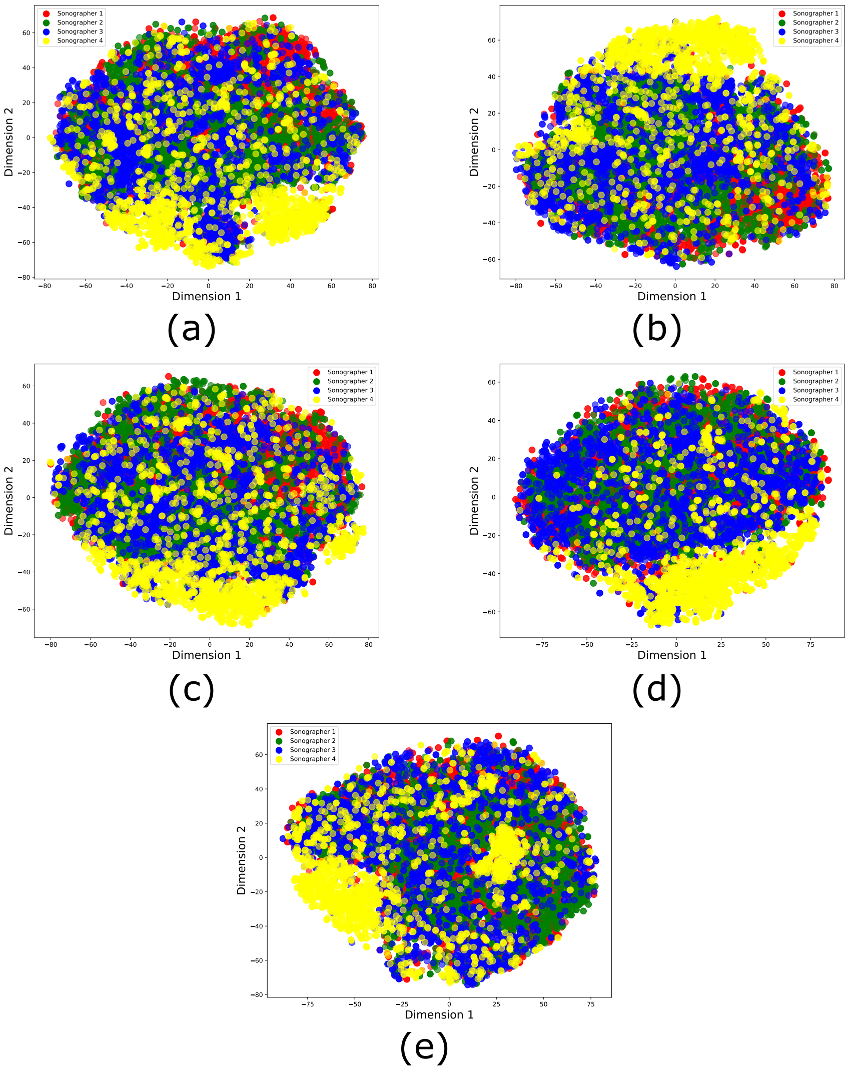


**Fig. S8**: **t-SNE coloured by sonographer.** (a) Encoder trained for B pattern. (b) Encoder trained for C2 pattern. (c) Encoder trained for E/F pattern. (d) Encoder trained on sweep two video. (e) Encoder trained on sweep three video.

#### S.5 Precision-recall curves

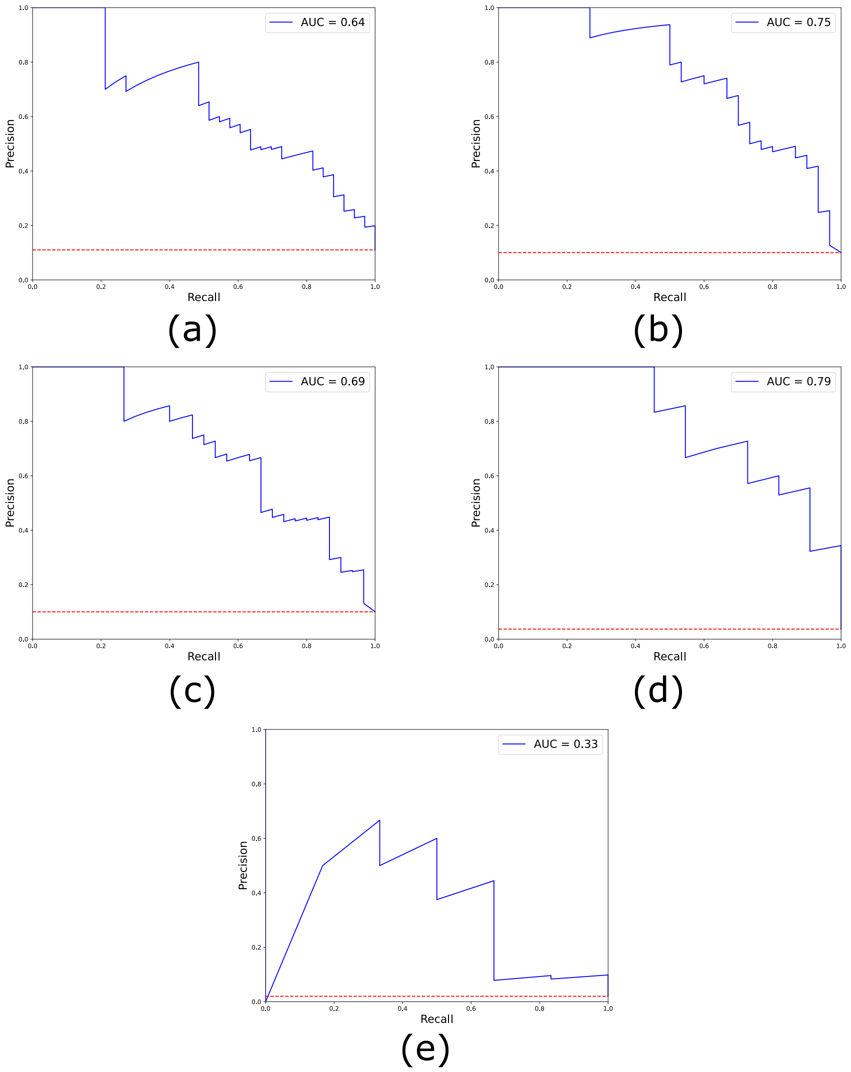


**Fig. S9**: **Precision-recall curves.** Precision-recall curves with AUC-PR indicated for each pattern of fibrosis. (a) B. (b) C1. (c) C2. (d) D. (e) E/F.

#### S.6 Participants with multiple patterns

**Table S5: Quantitative metrics among participants with different numbers of patterns.** Evaluated using the model at maximum F1-score.

| **Participants with 1 pattern** | | | | | | |
|---|---|---|---|---|---|---|
| | B | C1 | C2 | D | E/F | No Fibrosis |
| Accuracy | 0.64 | 0.84 | 0.88 | 0.96 | 0.96 | 0.64 |
| F1-score | 0.7692 | 0.0 | 0.0 | 0.6667 | 0.0 | 0.0 |
| Sensitivity | 0.625 | 0.0 | 0.0 | 1.0 | 0.0 | 0.0 |
| Specificity | 1.0 | 0.84 | 0.88 | 0.9583 | 0.96 | 0.64 |
| PPV | 1.0 | 0.0 | 0.0 | 0.5 | 0.0 | 0.0 |
| NPV | 0.1 | 1.0 | 1.0 | 1.0 | 1.0 | 1.0 |
| Prevalence | 0.96 | 0.0 | 0.0 | 0.04 | 0.0 | 0.0 |
| Count | 24 | 0 | 0 | 1 | 0 | 0 |
| **Participants with 2 patterns** | | | | | | |
| | B | C1 | C2 | D | E/F | No Fibrosis |
| Accuracy | 0.6923 | 0.6154 | 0.4615 | 0.9231 | 0.9231 | 0.6923 |
| F1-score | 0.3333 | 0.7368 | 0.6316 | 0.6667 | 0.0 | 0.0 |
| Sensitivity | 1.0 | 0.5833 | 0.5 | 1.0 | 0.0 | 0.0 |
| Specificity | 0.6667 | 1.0 | 0.0 | 0.9167 | 0.9231 | 0.6923 |
| PPV | 0.2 | 1.0 | 0.8571 | 0.5 | 0.0 | 0.0 |
| NPV | 1.0 | 0.1667 | 0.0 | 1.0 | 1.0 | 1.0 |
| Prevalence | 0.0769 | 0.9231 | 0.9231 | 0.0769 | 0.0 | 0.0 |
| Count | 1 | 12 | 12 | 1 | 0 | 0 |
| **Participants with 3 patterns** | | | | | | |
| | B | C1 | C2 | D | E/F | No Fibrosis |
| Accuracy | 0.7857 | 0.7143 | 0.7143 | 0.9286 | 0.7857 | 0.8571 |
| F1-score | 0.7273 | 0.8333 | 0.8333 | 0.9091 | 0.4 | 0.0 |
| Sensitivity | 0.8 | 0.7143 | 0.7143 | 0.8333 | 0.3333 | 0.0 |
| Specificity | 0.7778 | 0.0 | 0.0 | 1.0 | 0.9091 | 0.8571 |
| PPV | 0.6667 | 1.0 | 1.0 | 1.0 | 0.5 | 0.0 |
| NPV | 0.875 | 0.0 | 0.0 | 0.8889 | 0.8333 | 1.0 |
| Prevalence | 0.3571 | 1.0 | 1.0 | 0.4286 | 0.2143 | 0.0 |
| Count | 5 | 14 | 14 | 6 | 3 | 0 |

**Table S6**: **Quantitative metrics among participants with different numbers of patterns.** Evaluated using the model at 90% sensitivity.

| **Participants with 1 pattern** | | | | | | |
|---|---|---|---|---|---|---|
| | B | C1 | C2 | D | E/F | No Fibrosis |
| Accuracy | 0.88 | 0.6 | 0.36 | 0.96 | 0.6 | 0.88 |
| F1-score | 0.9333 | 0.0 | 0.0 | 0.6667 | 0.0 | 0.0 |
| Sensitivity | 0.875 | 0.0 | 0.0 | 1.0 | 0.0 | 0.0 |
| Specificity | 1.0 | 0.6 | 0.36 | 0.9583 | 0.6 | 0.88 |
| PPV | 1.0 | 0.0 | 0.0 | 0.5 | 0.0 | 0.0 |
| NPV | 0.25 | 1.0 | 1.0 | 1.0 | 1.0 | 1.0 |
| Prevalence | 0.96 | 0.0 | 0.0 | 0.04 | 0.0 | 0.0 |
| Count | 24 | 0 | 0 | 1 | 0 | 0 |
| **Participants with 2 patterns** | | | | | | |
| | B | C1 | C2 | D | E/F | No Fibrosis |
| Accuracy | 0.3077 | 0.8462 | 0.7692 | 0.9231 | 0.5385 | 0.8462 |
| F1-score | 0.1818 | 0.9091 | 0.8696 | 0.6667 | 0.0 | 0.0 |
| Sensitivity | 1.0 | 0.8333 | 0.8333 | 1.0 | 0.0 | 0.0 |
| Specificity | 0.25 | 1.0 | 0.0 | 0.9167 | 0.5385 | 0.8462 |
| PPV | 0.1 | 1.0 | 0.9091 | 0.5 | 0.0 | 0.0 |
| NPV | 1.0 | 0.3333 | 0.0 | 1.0 | 1.0 | 1.0 |
| Prevalence | 0.0769 | 0.9231 | 0.9231 | 0.0769 | 0.0 | 0.0 |
| Count | 1 | 12 | 12 | 1 | 0 | 0 |
| **Participants with 3 patterns** | | | | | | |
| | B | C1 | C2 | D | E/F | No Fibrosis |
| Accuracy | 0.4286 | 0.9286 | 0.9286 | 0.9286 | 0.5 | 1.0 |
| F1-score | 0.5556 | 0.963 | 0.963 | 0.9091 | 0.4615 | 0.0 |
| Sensitivity | 1.0 | 0.9286 | 0.9286 | 0.8333 | 1.0 | 0.0 |
| Specificity | 0.1111 | 0.0 | 0.0 | 1.0 | 0.3636 | 1.0 |
| PPV | 0.3846 | 1.0 | 1.0 | 1.0 | 0.3 | 0.0 |
| NPV | 1.0 | 0.0 | 0.0 | 0.8889 | 1.0 | 1.0 |
| Prevalence | 0.3571 | 1.0 | 1.0 | 0.4286 | 0.2143 | 0.0 |
| Count | 5 | 14 | 14 | 6 | 3 | 0 |

#### S.7 Results broken down by age and sex

**Table S7: Quantitative metrics for adults and children, evaluated at best F1-score.** Adults were defined as participants aged $\geq$ 15 at the time of ultrasound examination, and children were defined as participants aged <15 at the time of ultrasound examination.

| **Adults** | | | | | | |
|---|---|---|---|---|---|---|
| | B | C1 | C2 | D | E/F | No Fibrosis |
| Accuracy | 0.8773 | 0.9202 | 0.9018 | 0.9755 | 0.9693 | 0.8528 |
| F1-score | 0.4444 | 0.7547 | 0.7037 | 0.8333 | 0.5455 | 0.904 |
| Sensitivity | 0.5 | 0.8 | 0.7917 | 0.9091 | 0.5 | 0.904 |
| Specificity | 0.9184 | 0.942 | 0.9209 | 0.9803 | 0.9873 | 0.6842 |
| PPV | 0.4 | 0.7143 | 0.6333 | 0.7692 | 0.6 | 0.904 |
| NPV | 0.9441 | 0.963 | 0.9624 | 0.9933 | 0.981 | 0.6842 |
| Prevalence | 0.0982 | 0.1534 | 0.1472 | 0.0675 | 0.0368 | 0.7669 |
| Count | 16 | 25 | 24 | 11 | 6 | 125 |
| **Children** | | | | | | |
| | B | C1 | C2 | D | E/F | No Fibrosis |
| Accuracy | 0.9706 | 0.9706 | 0.9632 | 1.0 | 0.9926 | 0.9632 |
| F1-score | 0.8824 | 0.3333 | 0.2857 | 0.0 | 0.0 | 0.9789 |
| Sensitivity | 0.8824 | 0.2 | 0.1667 | 0.0 | 0.0 | 0.9831 |
| Specificity | 0.9832 | 1.0 | 1.0 | 1.0 | 0.9926 | 0.8333 |
| PPV | 0.8824 | 1.0 | 1.0 | 0.0 | 0.0 | 0.9748 |
| NPV | 0.9832 | 0.9704 | 0.963 | 1.0 | 1.0 | 0.8824 |
| Prevalence | 0.125 | 0.0368 | 0.0441 | 0.0 | 0.0 | 0.8676 |
| Count | 17 | 5 | 6 | 0 | 0 | 118 |

**Table S8**: **Quantitative metrics for adults and children, evaluated at 90% sensitivity.** Adults were defined as participants aged $\geq$ 15 at the time of ultrasound examination, and children were defined as participants aged $<$15 at the time of ultrasound examination.

| **Adults** | | | | | | |
|---|---|---|---|---|---|---|
| | B | C1 | C2 | D | E/F | No Fibrosis |
| Accuracy | 0.6748 | 0.8405 | 0.773 | 0.9755 | 0.8282 | 0.7423 |
| F1-score | 0.3291 | 0.6486 | 0.5542 | 0.8333 | 0.3 | 0.8056 |
| Sensitivity | 0.8125 | 0.96 | 0.9583 | 0.9091 | 1.0 | 0.696 |
| Specificity | 0.6599 | 0.8188 | 0.741 | 0.9803 | 0.8217 | 0.8947 |
| PPV | 0.2063 | 0.4898 | 0.3898 | 0.7692 | 0.1765 | 0.956 |
| NPV | 0.97 | 0.9912 | 0.9904 | 0.9933 | 1.0 | 0.4722 |
| Prevalence | 0.0982 | 0.1534 | 0.1472 | 0.0675 | 0.0368 | 0.7669 |
| Count | 16 | 25 | 24 | 11 | 6 | 125 |
| **Children** | | | | | | |
| | B | C1 | C2 | D | E/F | No Fibrosis |
| Accuracy | 0.8676 | 0.9338 | 0.8824 | 1.0 | 0.9338 | 0.8603 |
| F1-score | 0.6538 | 0.4 | 0.3333 | 0.0 | 0.0 | 0.9132 |
| Sensitivity | 1.0 | 0.6 | 0.6667 | 0.0 | 0.0 | 0.8475 |
| Specificity | 0.8487 | 0.9466 | 0.8923 | 1.0 | 0.9338 | 0.9444 |
| PPV | 0.4857 | 0.3 | 0.2222 | 0.0 | 0.0 | 0.9901 |
| NPV | 1.0 | 0.9841 | 0.9831 | 1.0 | 1.0 | 0.4857 |
| Prevalence | 0.125 | 0.0368 | 0.0441 | 0.0 | 0.0 | 0.8676 |
| Count | 17 | 5 | 6 | 0 | 0 | 118 |

**Table S9**: **Quantitative metrics for male and female participants, evaluated at best F1-score.**

| **Male** | | | | | | |
|---|---|---|---|---|---|---|
| | B | C1 | C2 | D | E/F | No Fibrosis |
| Accuracy | 0.9562 | 0.9343 | 0.9416 | 0.9781 | 0.9781 | 0.9416 |
| F1-score | 0.8235 | 0.7429 | 0.7647 | 0.8421 | 0.6667 | 0.963 |
| Sensitivity | 0.9333 | 0.7647 | 0.7647 | 0.8889 | 0.6 | 0.963 |
| Specificity | 0.959 | 0.9583 | 0.9667 | 0.9844 | 0.9924 | 0.8621 |
| PPV | 0.7368 | 0.7222 | 0.7647 | 0.8 | 0.75 | 0.963 |
| NPV | 0.9915 | 0.9664 | 0.9667 | 0.9921 | 0.985 | 0.8621 |
| Prevalence | 0.1095 | 0.1241 | 0.1241 | 0.0657 | 0.0365 | 0.7883 |
| Count | 15 | 17 | 17 | 9 | 5 | 108 |
| **Female** | | | | | | |
| | B | C1 | C2 | D | E/F | No Fibrosis |
| Accuracy | 0.8889 | 0.9506 | 0.9198 | 0.9938 | 0.9815 | 0.8704 |
| F1-score | 0.5 | 0.6667 | 0.5185 | 0.8 | 0.0 | 0.9225 |
| Sensitivity | 0.5 | 0.6154 | 0.5385 | 1.0 | 0.0 | 0.9259 |
| Specificity | 0.9375 | 0.9799 | 0.953 | 0.9938 | 0.9876 | 0.5926 |
| PPV | 0.5 | 0.7273 | 0.5 | 0.6667 | 0.0 | 0.9191 |
| NPV | 0.9375 | 0.9669 | 0.9595 | 1.0 | 0.9937 | 0.6154 |
| Prevalence | 0.1111 | 0.0802 | 0.0802 | 0.0123 | 0.0062 | 0.8333 |
| Count | 18 | 13 | 13 | 2 | 1 | 135 |

**Table S10**: **Quantitative metrics for male and female participants, evaluated at 90% sensitivity.**

| Male | | | | | | |
|---|---|---|---|---|---|---|
| | B | C1 | C2 | D | E/F | No Fibrosis |
| Accuracy | 0.7445 | 0.8686 | 0.8248 | 0.9781 | 0.854 | 0.8175 |
| F1-score | 0.4615 | 0.64 | 0.5714 | 0.8421 | 0.3333 | 0.8691 |
| Sensitivity | 1.0 | 0.9412 | 0.9412 | 0.8889 | 1.0 | 0.7685 |
| Specificity | 0.7131 | 0.8583 | 0.8083 | 0.9844 | 0.8485 | 1.0 |
| PPV | 0.3 | 0.4848 | 0.4103 | 0.8 | 0.2 | 1.0 |
| NPV | 1.0 | 0.9904 | 0.9898 | 0.9921 | 1.0 | 0.537 |
| Prevalence | 0.1095 | 0.1241 | 0.1241 | 0.0657 | 0.0365 | 0.7883 |
| Count | 15 | 17 | 17 | 9 | 5 | 108 |
| **Female** | | | | | | |
| | B | C1 | C2 | D | E/F | No Fibrosis |
| Accuracy | 0.7778 | 0.8951 | 0.821 | 0.9938 | 0.8951 | 0.7778 |
| F1-score | 0.4545 | 0.5641 | 0.4314 | 0.8 | 0.1053 | 0.8525 |
| Sensitivity | 0.8333 | 0.8462 | 0.8462 | 1.0 | 1.0 | 0.7704 |
| Specificity | 0.7708 | 0.8993 | 0.8188 | 0.9938 | 0.8944 | 0.8148 |
| PPV | 0.3125 | 0.4231 | 0.2895 | 0.6667 | 0.0556 | 0.9541 |
| NPV | 0.9737 | 0.9853 | 0.9839 | 1.0 | 1.0 | 0.4151 |
| Prevalence | 0.1111 | 0.0802 | 0.0802 | 0.0123 | 0.0062 | 0.8333 |
| Count | 18 | 13 | 13 | 2 | 1 | 135 |

**Table S11: Summary of interference prevalence and clinical confidence in 2024 test set.**

| Characteristic | Percentage |
|---|---|
| *Interference* | |
| Gas | 17.2% (10/58) |
| Breathing/movement | 6.9% (4/58) |
| Dehydration | 3.4% (2/58) |
| Fat | 1.7% (1/58) |
| Bone | 0% (0/58) |
| Calcification | 0% (0/58) |
| *Clinical confidence* | |
| High | 55.2% (32/58) |
| Medium | 32.8% (19/58) |
| Low | 12.0% (7/58) |

#### S.8 Errors in videos with interference or comorbidities

**Table S12: Summary of interference prevalence, distribution of clinical confidence and prevalence of ultrasound findings in errors in the 2024 test set.**

| Characteristic | B | C1 | C2 | D | E/F |
|---|---|---|---|---|---|
| *Interference* | | | | | |
| Gas | 5/15 | 2/9 | 1/7 | 0/1 | 0/1 |
| Breathing/movement | 3/15 | 1/9 | 0/7 | 0/1 | 0/1 |
| Dehydration | 1/15 | 0/9 | 0/7 | 0/1 | 0/1 |
| Fat | 1/15 | 1/9 | 0/7 | 0/1 | 0/1 |
| *Clinical confidence* | | | | | |
| High | 9/15 | 8/9 | 6/7 | 1/1 | 1/1 |
| Moderate | 4/15 | 1/9 | 1/7 | 0/1 | 0/1 |
| Low | 2/15 | 0/9 | 0/7 | 0/1 | 0/1 |

#### S.9 Sensitivity analyses

**Table S13: Performance of different setups for predicting probabilities.**

| **Overall** | | | | | | |
|---|---|---|---|---|---|---|
| | B | C1 | C2 | D | E/F | |
| *Maximum* | | | | | | |
| AUROC | 0.8757 | 0.9135 | 0.9024 | 0.9752 | 0.8834 | |
| AUC-PR | 0.585 | 0.6761 | 0.6231 | 0.5868 | 0.295 | |
| AUC-PR:prevalence | 5.3002 | 6.7389 | 6.2101 | 15.9504 | 14.6988 | |
| *Mean* | | | | | | |
| AUROC | 0.8978 | 0.9281 | 0.9099 | 0.9815 | 0.8879 | |
| AUC-PR | 0.5952 | 0.7424 | 0.6774 | 0.7277 | 0.3534 | |
| AUC-PR:prevalence | 5.3925 | 7.3994 | 6.7518 | 19.7809 | 17.6121 | |
| *Clips* | | | | | | |
| AUROC | 0.9156 | 0.9314 | 0.9141 | 0.9867 | 0.9369 | |
| AUC-PR | 0.6448 | 0.7529 | 0.6893 | 0.7912 | 0.3285 | |
| AUC-PR:prevalence | 5.842 | 7.5034 | 6.8699 | 21.5072 | 16.3719 | |
| **At maximum F1-score** | | | | | | |
| | B | C1 | C2 | D | E/F | No Fibrosis |
| *Maximum* | | | | | | |
| Accuracy | 0.9097 | 0.9331 | 0.9064 | 0.9498 | 0.9833 | 0.8696 |
| F1-score | 0.5846 | 0.6 | 0.6316 | 0.5714 | 0.5455 | 0.9172 |
| Sensitivity | 0.5758 | 0.5 | 0.8 | 0.9091 | 0.5 | 0.8889 |
| Specificity | 0.9511 | 0.9814 | 0.9182 | 0.9514 | 0.9932 | 0.7857 |
| PPV | 0.5938 | 0.75 | 0.5217 | 0.4167 | 0.6 | 0.9474 |
| NPV | 0.9476 | 0.9462 | 0.9763 | 0.9964 | 0.9898 | 0.6197 |
| *Mean* | | | | | | |
| Accuracy | 0.913 | 0.9365 | 0.9197 | 0.9766 | 0.9866 | 0.8829 |
| F1-score | 0.5667 | 0.678 | 0.625 | 0.6957 | 0.6 | 0.9281 |
| Sensitivity | 0.5152 | 0.6667 | 0.6667 | 0.7273 | 0.5 | 0.93 |
| Specificity | 0.9624 | 0.9665 | 0.948 | 0.9861 | 0.9966 | 0.6786 |
| PPV | 0.6296 | 0.6897 | 0.5882 | 0.6667 | 0.75 | 0.9262 |
| NPV | 0.9412 | 0.963 | 0.9623 | 0.9895 | 0.9898 | 0.6909 |

| | | | | | | |
|---|---|---|---|---|---|---|
| *Clips* | | | | | | |
| Accuracy | 0.9197 | 0.9431 | 0.9298 | 0.9866 | 0.9799 | 0.9030 |
| F1-score | 0.6571 | 0.7119 | 0.6557 | 0.8333 | 0.5000 | 0.9405 |
| Sensitivity | 0.6970 | 0.7000 | 0.6667 | 0.9091 | 0.5000 | 0.9424 |
| Specificity | 0.9474 | 0.9703 | 0.9591 | 0.9896 | 0.9898 | 0.7321 |
| PPV | 0.6216 | 0.7241 | 0.6452 | 0.7692 | 0.5000 | 0.9385 |
| NPV | 0.9618 | 0.9667 | 0.9627 | 0.9965 | 0.9898 | 0.7455 |
| **At 90% sensitivity** | | | | | | |
| | B | C1 | C2 | D | E/F | No Fibrosis |
| *Maximum* | | | | | | |
| Accuracy | 0.5987 | 0.8696 | 0.7191 | 0.9498 | 0.0201 | 0.1873 |
| F1-score | 0.3333 | 0.5806 | 0.3913 | 0.5714 | 0.0393 | 0.0 |
| Sensitivity | 0.9091 | 0.9 | 0.9 | 0.9091 | 1.0 | 0.0 |
| Specificity | 0.5602 | 0.8662 | 0.6989 | 0.9514 | 0.0 | 1.0 |
| PPV | 0.2041 | 0.4286 | 0.25 | 0.4167 | 0.0201 | 0.0 |
| NPV | 0.9803 | 0.9873 | 0.9843 | 0.9964 | 0.0 | 0.1873 |
| *Mean* | | | | | | |
| Accuracy | 0.699 | 0.8595 | 0.7692 | 0.9565 | 0.0201 | 0.1873 |
| F1-score | 0.4 | 0.5625 | 0.439 | 0.6061 | 0.0393 | 0.0 |
| Sensitivity | 0.9091 | 0.9 | 0.9 | 0.9091 | 1.0 | 0.0 |
| Specificity | 0.6729 | 0.855 | 0.7546 | 0.9583 | 0.0 | 1.0 |
| PPV | 0.2564 | 0.4091 | 0.2903 | 0.4545 | 0.0201 | 0.0 |
| NPV | 0.9835 | 0.9871 | 0.9854 | 0.9964 | 0.0 | 0.1873 |
| *Clips* | | | | | | |
| Accuracy | 0.7659 | 0.8829 | 0.7793 | 0.9699 | 0.8127 | 0.7492 |
| F1-score | 0.4615 | 0.6067 | 0.45 | 0.6897 | 0.1765 | 0.8201 |
| Sensitivity | 0.9091 | 0.9 | 0.9 | 0.9091 | 1.0 | 0.7037 |
| Specificity | 0.7481 | 0.881 | 0.7658 | 0.9722 | 0.8089 | 0.9464 |
| PPV | 0.3093 | 0.4576 | 0.3 | 0.5556 | 0.0968 | 0.9828 |
| NPV | 0.9851 | 0.9875 | 0.9856 | 0.9964 | 1.0 | 0.424 |

**Table S14**: **Performance of model with one district held out.** A model was trained on two districts (Pakwach and Buliisa) and tested on a third (Mayuge).

| **Overall** | | | | | | |
|---|---|---|---|---|---|---|
| | B | C1 | C2 | D | E/F | |
| AUROC | 0.8768 | 0.9524 | 0.956 | 0.9612 | 0.932 | |
| AUC-PR | 0.481 | 0.6431 | 0.6836 | 0.6909 | 0.2999 | |
| AUC-PR:prevalence | 6.9816 | 9.0744 | 9.1391 | 15.9529 | 30.469 | |
| **At maximum F1-score** | | | | | | |
| | B | C1 | C2 | D | E/F | No Fibrosis |
| Accuracy | 0.9055 | 0.9409 | 0.9409 | 0.9764 | 0.9921 | 0.9134 |
| F1-score | 0.5102 | 0.625 | 0.6667 | 0.6667 | 0.5 | 0.9486 |
| Sensitivity | 0.7143 | 0.6944 | 0.7895 | 0.5455 | 0.4 | 0.9355 |
| Specificity | 0.9197 | 0.9597 | 0.9532 | 0.9959 | 0.998 | 0.7838 |
| PPV | 0.3968 | 0.5682 | 0.5769 | 0.8571 | 0.6667 | 0.9621 |
| NPV | 0.9775 | 0.9763 | 0.9825 | 0.9798 | 0.9941 | 0.6744 |
| **At 90% sensitivity** | | | | | | |
| | B | C1 | C2 | D | E/F | No Fibrosis |
| Accuracy | 0.6949 | 0.8504 | 0.8504 | 0.876 | 0.878 | 0.7579 |
| F1-score | 0.2922 | 0.4648 | 0.4795 | 0.3883 | 0.1389 | 0.8362 |
| Sensitivity | 0.9143 | 0.9167 | 0.9211 | 0.9091 | 1.0 | 0.7235 |
| Specificity | 0.6786 | 0.8453 | 0.8447 | 0.8745 | 0.8767 | 0.9595 |
| PPV | 0.1739 | 0.3113 | 0.3241 | 0.2469 | 0.0746 | 0.9905 |
| NPV | 0.9907 | 0.9925 | 0.9925 | 0.9953 | 1.0 | 0.3717 |
